# Effective correction of extreme capacitive artifacts in TMS-EEG via windowed detrending

**DOI:** 10.64898/2025.12.03.25341368

**Authors:** A. A. Vergani, G. Peroni, C. Bruscagli, A. Lionti, F. Brandizzi, M. Lenge, G. Salvestrini, D. Jiménez - Jiménez, S. Casarotto, M. Rosanova, A. Mazzoni, R. Guerrini, A. Grippo, S. Balestrini

## Abstract

**Background:** Transcranial Magnetic Stimulation (TMS) combined with electroencephalography (EEG) is a tool for investigating non-invasively cortical excitability and connectivity. However, this technique is susceptible to artifacts that can obscure genuine neural activity, making signal correction a critical component of data processing. Among these, capacitive artifacts pose a significant challenge, as they hinder the accurate interpretation of TMS-evoked potentials (TEPs). These artifacts can often be mitigated online through optimized recording settings and experimental procedures, but in the most challenging cases they must be removed offline to ensure accurate reconstruction of TEPs.

**Methods:** To correct capacitive artifacts, we tested detrending methods based on both windowed and non-windowed approaches. Non-windowed methods operate on the full EEG epoch, while windowed approaches segment the signal into rise and decay phases of the post-stimulus signal, which correspond to the electrode-related charging and discharging effects triggered by the stimulation. We applied these methods on several datasets from two centers acquired with different hardware settings. We controlled capacitive artifacts and benchmarked detrending algorithm performance against a cleaning strategy based on Independent Component Analysis (ICA).

**Results:** Capacitive artifacts varied from mild to extreme, depending on the amplitude and characteristic time constants of the capacitive model. ICA was effective in mild cases but failed to adequately correct moderate to severe artifacts. In contrast, model-based detrending approaches, particularly windowed methods, proved more effective in managing extreme artifacts, as segmented fitting facilitates parameter identification. Among these, windowed polynomial detrending showed a slight advantage due to its adaptability to complex artifact shapes.

**Conclusions:** While the most effective way to minimize TMS-related capacitive artifacts is to use optimal TMS-compatible hardware together with rigorous online prevention and monitoring during acquisition, reliable offline correction methods remain essential when ideal recording conditions cannot be guaranteed. In this context, windowed detrending provides a robust offline strategy that improves the reliability of TEP interpretation, particularly in settings where hardware optimization is not feasible.

**Highlights:**

- Capacitive artifacts in TMS-EEG recordings vary in severity and can compromise the interpretation of TMS-evoked potentials (TEPs)
- Windowed detrending outperforms ICA and non-windowed models in correcting extreme capacitive artifacts and restoring the physiological 1/f spectral profile.
- Advanced signal correction methods enhance the reliability of TMS-EEG recordings, particularly when artifacts are source-related or when affected channels cannot be excluded.

## 1. Introduction

Transcranial magnetic stimulation (TMS) combined with electroencephalography (EEG) enables non-invasive assessment of key cortical properties through TMS-evoked potentials (TEPs) (Kuhn et al., 2025; She et al., 2024). However, obtaining clean TEPs is challenging due to numerous artifacts introduced by the TMS pulse delivery (Bertazzoli et al., 2021; Brancaccio et al., 2024; Hernandez-Pavon et al., 2022; Mutanen et al., 2022).

TMS-EEG artifacts are generally classified as either physiological or non-physiological artifacts (Miniussi & Thut, 2010; Varone et al., 2021; Vernet & Thut, 2014). Physiological artifacts arise from the brain or body’s response to stimulation and can obscure neural signals. These include eye blinks and movements, which generate large frontal deflections due to the startle response or spontaneous ocular activity; cranial muscle artifacts producing biphasic deflections from scalp and facial muscle contractions following direct nerve stimulation (Ma et al., 2012; Mutanen et al., 2016); auditory evoked potentials, arising from the coil click, contaminating late TEP components (Conde et al., 2019; Siebner et al., 2022); somatosensory responses, caused by peripheral nerve activation due to coil vibrations or peripheral nerve activation (but see Mancuso et al., 2023; 10.1016/j.neubiorev.2023.105434). Non-physiological artifacts, in contrast, may result from direct interactions between the TMS pulse and the EEG recording hardware. These include the pulse artifact, a high-amplitude disturbance that may saturate amplifiers; the recharge artifact, caused by capacitor recharge within the stimulator; movement artifacts, due to electrode displacement from coil vibrations or muscle contractions (Burbank & Webster, 1978); eddy currents, especially in non-slitted electrodes, producing transient voltage shifts (Ruddy et al., 2018); thermal artifacts (Johnson, 1928; Nyquist, 1928); electrode polarization artifacts, caused by charge buildup at the electrode-electrolyte interface, leading to slowly decaying drifts known as ‘capacitive artifacts’ or ‘decay artifacts’ (Casarotto et al., 2022; Ilmoniemi et al., 2015; Ilmoniemi & Kičić, 2010; Litvak et al., 2007; Rogasch et al., 2017; Vernet & Thut, 2014). Extreme capacitive artifacts may present with a sharp initial voltage deflection (extreme rise) followed by a prolonged, nonlinear return to baseline (extreme decay) (J. Virtanen et al., 1999).

Several hardware strategies can mitigate these artifacts, including the use of slitted electrodes (C-ring shape) to minimise eddy currents; Ag/AgCl electrodes for lower impedance; and maintaining physical separation between the coil and electrodes (Brancaccio et al., 2024; Casarotto et al., 2022; Mutanen et al., 2020; Stango et al., 2025; Wu et al., 2018). Nevertheless, the specific combination of stimulator, EEG cap, and amplifier can introduce strong capacitive artifacts, whose temporal profile often dominates the recorded signal. However, these are sometimes insufficient in cases of severe non-linear capacitive behavior and excluding contaminated channels is not ideal, as they are typically closest to the stimulation site for capturing the neural activity in the underlying cortical region. Therefore, technical strategies capable of handling these signals during pre-processing without discarding relevant signals are essential.

Known artifact correction tools, such as ARTIST (Wu et al., 2018), TESA (Mutanen et al., 2020), and SOUND/SSP-SIR (Mutanen et al., 2016, 2020, 2022), often rely on Independent Component Analysis (ICA) or detrending-based strategies. ICA can be effective when the capacitive component is sufficiently distinct and separable; however, the large amplitude and sharp transients of extreme capacitive artifacts can dominate the signal, impairing ICA’s ability to isolate other components (Atti et al., 2024; Djuwari et al., 2005). In contrast, detrending methods, including polynomial-based approaches (demeaning, linear and non-linear detrending) and physiologically inspired methods that model the artifact using exponential functions or power law functions, are better suited for handling slow, high-amplitude drifts (Freche et al., 2018; Lentka & Smulko, 2019; Xu et al., 2009). These methods subtract estimated artifact dynamics from the EEG trace to restore physiological signal morphology. However, under extreme non- linearities, global model fitting becomes computationally challenging (Transtrum et al., 2010), and parameter estimation may fail.

In this study, we tackle the correction of extreme capacitive artifacts using both exponential and polynomial detrending strategies. We propose that windowed approaches, which separate the signal into rise and decay phases, provide more robust correction than traditional non-windowed, full-epoch methods. This segmentation facilitates more accurate parameter estimation and better artifact removal, especially when the signal exhibits non-linear dynamics.

## 2 Materials and Methods

### 2.1 Conceptual framework for mitigating extreme capacitive artifacts using detrending algorithms

We defined extreme capacitive artifacts based on the parameter values of exponential rise and decay functions used to model the full capacitive dynamics of the EEG signal, without temporal segmentation. We labeled a configuration as ‘extreme’ if the model fitting algorithm used for detrending (i) failed to converge due to computational instability, or (ii) produced an inadequate fit to the artifact signal. These failures occurred particularly when the rise time constant was very small (tau rise, e.g., approximately of < 10ms), resulting in a near-instantaneous onset, combined with a large decay time constant (tau decay, e.g., if > approximately of 100ms) yielding a prolonged return to baseline (see Supporting Figure 1 A-B).

**Figure 1.**
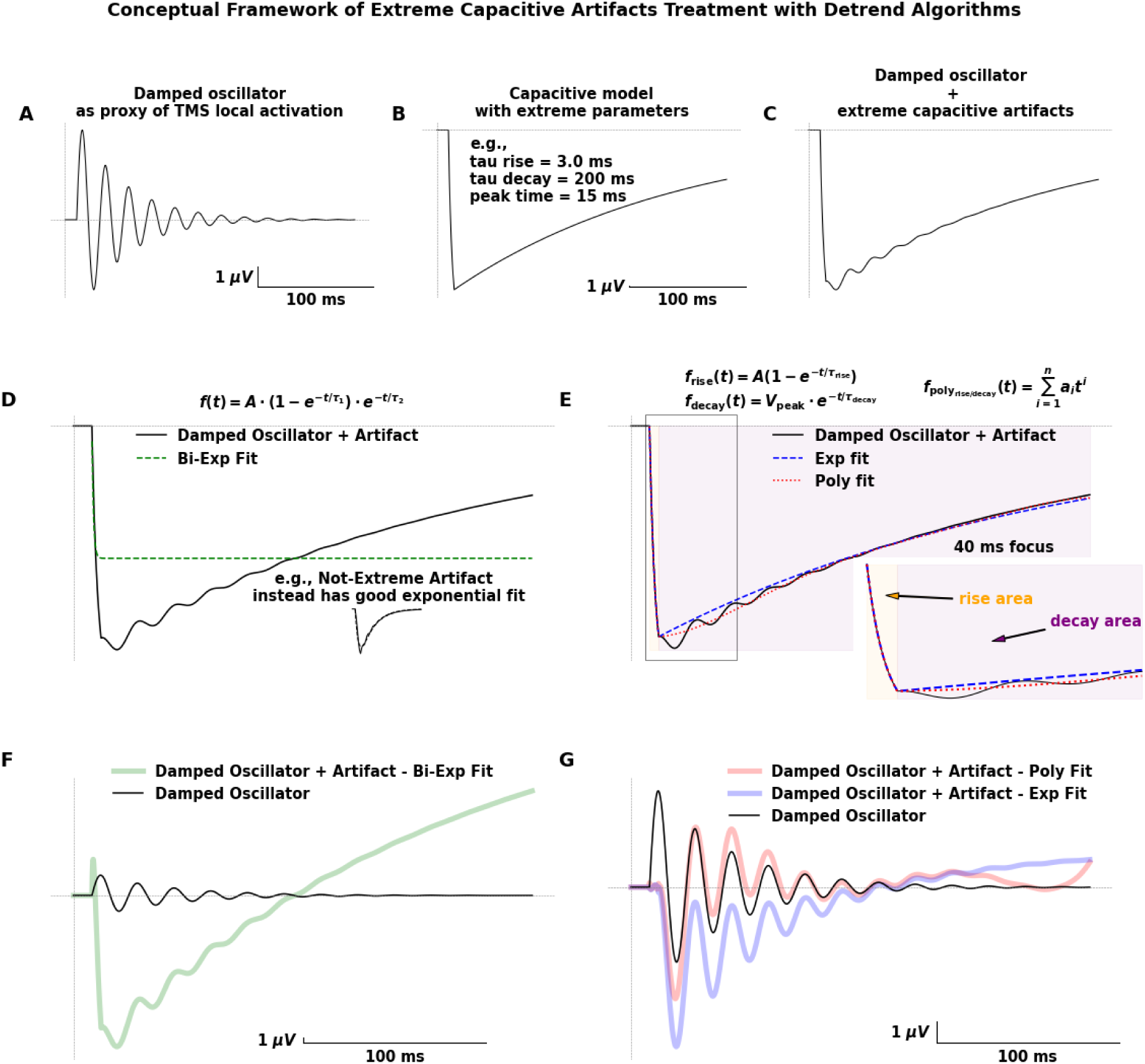
Framework for correcting extreme capacitive artifacts in TMS-EEG using windowed detrending approaches. **(A)** A damped oscillator simulates the neural response to transcranial magnetic stimulation (TMS). **(B)** Extreme capacitive artifacts are modeled using a physiologically plausible exponential rise (Tau Rise = 3 ms) and decay (Tau Decay = 200 ms), with a peak latency of 15 ms. **(C)** The resulting signal, combining neural response and artifact, exhibits strong capacitive contamination. **(D)** Non-windowed bi-exponential detrending fails to adequately capture the slow artifact dynamics, leading to poor correction (green fit). This approach may perform better on milder artifacts (inset). **(E)** Windowed detrending separates rise and decay phases and fits them independently using either exponential (blue) or polynomial (pink) functions within short temporal windows (e.g., 40 ms), improving model accuracy. **(F)** Subtraction of poor non-windowed fit leaves substantial residual artifacts, distorting the underlying neural response. **(G)** In contrast, windowed exponential and polynomial detrending more effectively isolate and remove the artifact, preserving oscillatory morphology. A more technical visualization is presented in Supporting Figure 1 (S-Fig1), where a broad family of simulated capacitive artifacts and their contaminating effects on a damped oscillator are illustrated, along with detailed results on model fitting and detrending performance with and without temporal windowing. Panel D/E/F share the same x/y axis scale.

We decomposed a simulated TEP with an extreme capacitive artifact into two components. The first is the true cortical response modeled as a damped oscillator (Figure 1A) (for the model equation refer to the paper by Serway and Jewett (Serway & Jewett, 2004)). The second is the capacitive distortion modeled via a sharp exponential rise and slow decay (Figure 1B). The combined signal is affected by a strong capacitive contamination (Figure 1C). Correcting such artificial perturbation involves reconstructing the capacitive waveform to be subtracted from the distorted signal, allowing recovery of the original damped oscillation. When fitting is performed on the entire epoch using a global (non-windowed) bi-exponential model, the extreme tau parameters often lead to failure in identifying a plausible artifact trend (Figure 1D). Instead, a windowed approach, which separates the signal at the artifact peak (the cusp) into rise and decay phases, enabled successful and independent fitting for each segment, producing parameters that capture the nonlinear trend aligned with both deflections (Figure 1E). We implemented and compared both exponential and polynomial fits (3rd order) for each phase. From the conceptual framework showcase, the results were broadly similar, the polynomial model showed slightly better performance, particularly in modeling the decay phase due to its greater adaptability to signal shape. Non-windowed detrending performed poorly in reconstructing the original neural oscillation (Figure 1F), whereas windowed detrending more accurately recovered the expected waveform with either exponential or polynomial fits (Figure 1G). For the detailed computational analysis of different cases of capacitive artefacts refer to Supporting Figure 1 C-F.

### 2.2 Participants and data collection

To identify TEP recordings affected by extreme capacitive artifacts, we tested two healthy (a Female with age range 25-30 years and a Male with age range 50-55 years) participants recording left premotor TEPs under various hardware configurations at the University of Milan, Department of Biomedical and Clinical Sciences. The study was approved by the local ethics committee (London South East Research Ethics Committee; REC reference: Cortical Excitability, 15/LO/1642). All participants gave written informed consent. We systematically varied electrodes (EasyCap 64- channel Standard BrainCap with passive C-slit sintered Ag/Ag-Cl electrodes, 10-20 system, or EBNeuroCap, custom 32- channel cap with passive C-slit Ag/Ag-Cl electrodes), amplifiers (Brainamp DC, Brain Products GmbH, Germany, DC- 1000 Hz bandwidth, 5000Hz sampling, or BE PLUS PRO, EB Neuro, Italy, 0.1-1000 Hz filtering, 4096 Hz sampling), stimulators (EBNeuroStim, STM9000 stimulator, butterfly coil 70mm - 90° with biphasic pulse setting, EB Neuro, Italy or Nexstim, Focal Bipulse 8-Coil stimulator, 50/70 mm diameter, 280 μs biphasic pulse, Nexstim Ltd., Finland). These combinations yielded the following datasets: D1: EasyCap + BrainAmp + Nexstim; D2: EasyCap + EBNeuro amplifier + Nexstim; D3: EasyCap + BrainAmp + EBNeuroStim; D4: EasyCap + EBNeuro amplifier + EBNeuroStim; D7: EBNeuroCap + BrainAmp + Nexstim.

We collected two additional datasets (D5, D6) at the Chalfont Centre for Epilepsy, (Buckinghamshire, UK), from already published work (D’Ambrosio et al., 2022). This study was approved by the local NHS research ethics committee (REF: 15/LO/1642). Here, TMS-EEG was recorded from two healthy participants delivering pulses with a figure of eight coil with 70-mm diameter driven by a monophasic stimulator Magstim 2002 (Whitland, UK) while recording was performed following international standards (Nuwer et al., 1998; Sinha et al., 2016), with an ActiCHamp 63-channel amplifier, and active TMS-compatible actiCAP electrodes (Brain Products GmbH, Germany) placed in the international 10–20 montage referenced to the forehead. The impedance of all electrodes was kept below 5 kΩ. EEG signals were band-pass filtered between 0.1 and 500<Hz and sampled at 5 kHz with 32-bit resolution. Real-time visualization via rt-TEP (Casarotto et al., 2022) was used to optimize stimulation parameters.

All datasets (D1-D7) were pre-processed, and those showing extreme capacitive artifacts were further analysed using both windowed and non-windowed detrending approaches.

### 2.3 TEP processing pipeline

#### 2.3.1 Pre-detrend phase

The following steps were performed before detrending. First, the TMS pulse artifact was removed by gap-filling method (Vafidis et al., 2019), replacing the lowest possible amount of contaminated EEG samples (from −2 to +5/+8 ms) with a mirrored version of the preceding baseline, then smoothing them using a 4 ms moving average (Casarotto et al., 2022; Hernandez-Pavon et al., 2022). Second, we applied broadband filtering between 0.1 Hz and 250 Hz using a 3rd order Butterworth IIR filter. Third, we used a notch FIR filter to remove 50 Hz line noise and its harmonics. Fourth, we used a separate 0.1-45 Hz Butterworth IIR filter for trial/channel rejection. Fifth, signals were segmented from −800 to +800 ms around the TMS pulse to identify and reject bad trials and channels. Sixth, we performed final epoching with broadband-filtered data. Finally, we downsampled the data to 1000 Hz, and re-referenced them to the average across channels.

#### 2.3.2 Detrend phase

The detrending phase involved a trial-wise investigation of anomalies caused by capacitive artifacts. When applying windowed detrending, we segmented the signal at the artifact peak into rise and decay phases by locating the extreme value that marks the transition point between them. We fitted each phase separately using either polynomial or exponential functions. Specifically, we preferred polynomial models over exponential ones for both rise and decay phases, as this formulation provided superior performance with the tested capacitive artifact models (see Figure 1 and Supporting Figure 1). For the rise phase, Lagrange polynomials of optimized degree were used to maximize fitting accuracy and ensure complete artifact removal. Once the rapid transient was fully captured in this phase, the decay phase was modeled using a fixed third-degree polynomial, selected to represent the slow nonlinear return to baseline while avoiding numerical instability and long-range overfitting typically associated with higher-order fits. The exact optimization of the decay polynomial degree is beyond the scope of this paper; however, low-order polynomials (from second to fifth degree) are generally sufficient to capture the nonlinear behavior of the decay phase. Since no ground-truth signal is available for validation, a third-degree polynomial was empirically chosen as a reasonable trade-off between flexibility and stability. This combined approach allows accurate modeling of the non-oscillatory trend without distorting the underlying oscillatory components, which might otherwise be attenuated by overly flexible models. However, for completeness, results using exponential fits within the windowed detrending approach are reported in Supporting Figure 2. In contrast, for non-windowed detrending, we employed a bi-exponential model to fit the entire signal, modelling capacitive charging and discharging dynamics. We performed exponential curve fitting using the *curve_fit* algorithm from the *scipy.optimize* module in Python for estimating equation parameters via non-linear least squared method (P. Virtanen et al., 2020)

**Figure 2:**
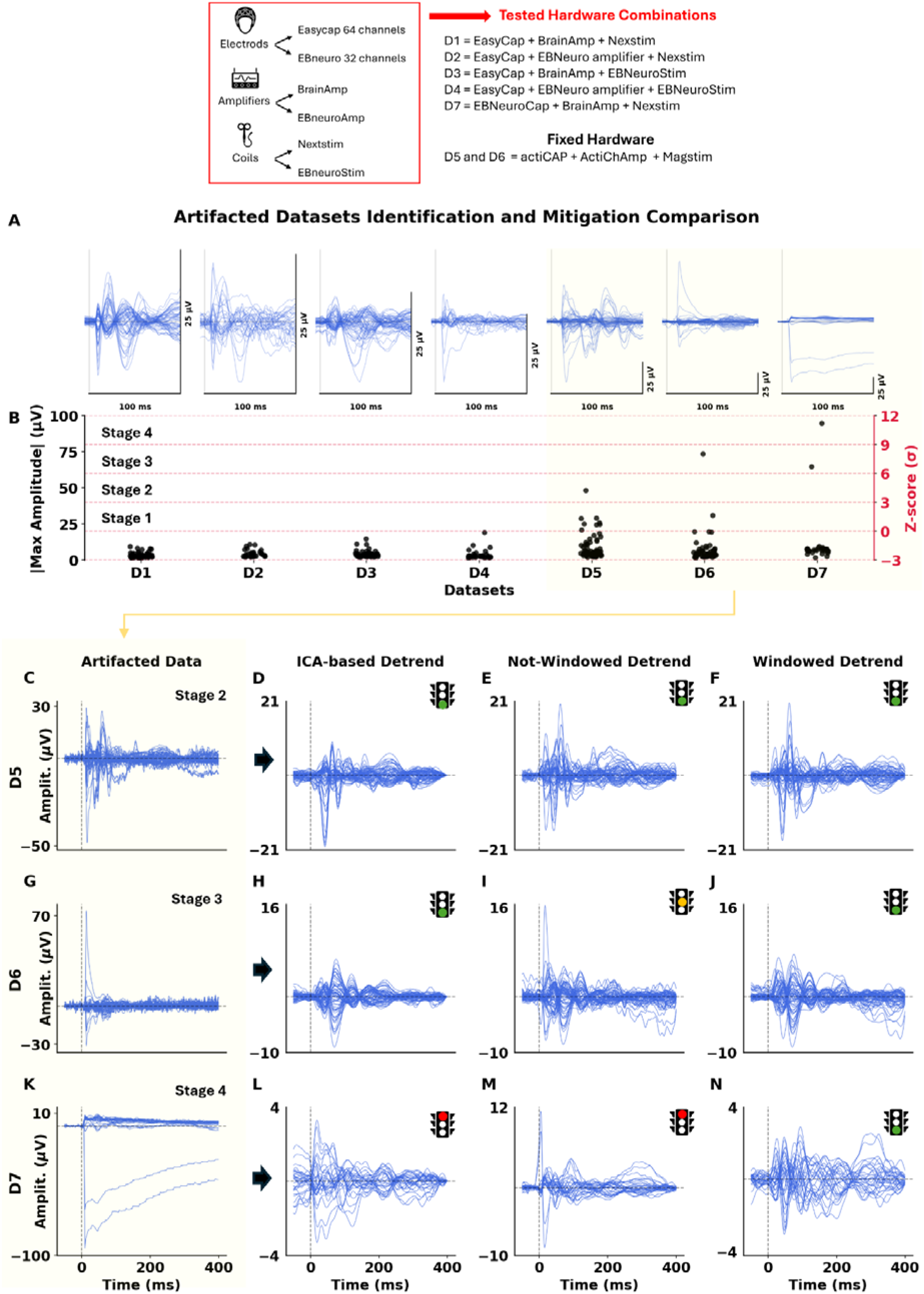
Identification and correction of capacitive artifacts across datasets. **(A)** Epoched EEG signals for each dataset (D1-D7), showing increasing artifact severity from left to right **(B)** Maximum channel amplitudes (left axis) and Z-scores (right axis) for each dataset, used to classify artifact severity into four stages (Stage 1-4). (**C, G, K**) Raw data for datasets D5 (Stage 2), D6 (Stage 3), and D7 (Stage 4). (**D, H, L**) ICA-based correction results. (**E, I, M**) Non-windowed bi-exponential detrending results, showing inconsistent performance depending on artifact severity. (**F, J, N**) Windowed polynomial detrending results, providing better artifact removal and morphology preservation, particularly effective in severe cases (Stage 3-4). A traffic-light indicator summarizes signal quality: green = acceptable, yellow = partially acceptable, red = inadequate. The color classification reflects a compromise between qualitative inspection, denoising performance (Figure 3) and amplitude correction (Supporting Figure 8). The windowed detrending method consistently outperforms other methods in artifact suppression and signal preservation, especially in Stage 3 and 4 cases. Polynomial fits were used for windowed detrending (see Methods); corresponding exponential-fit results are presented in Supporting Figure 3. The ICA components removed for the ICA-based detrend column are shown in Supporting Figure 4, 5 and 6. For a detailed analysis of trial-wise effects at Cz, refer to Supporting Figure 7.

As an alternative to non-windowed detrending, we applied ICA alone to isolate artifact-related components by visually inspecting the component time courses and excluding those clearly representing the scalp potential trend (an example of capacitive artifact ICA component is the decay trend in figure 6 in (Wu et al., 2018). All the ICA removed components are in the supporting section. After excluding these components, the signal was reconstructed for comparison with the detrending-based method. Visualizations in the figures were generated using data filtered between 1 and 30 Hz with a Butterworth IIR filter.

### 2.4. Denoising Estimation with Spectral Parameterization (FOOOF)

In neural time series, broadband noise and scale-free background activity are captured by the aperiodic component of the power spectrum, which reflects non-oscillatory, stochastic fluctuations often associated with neural noise or synaptic background activity (Voytek et al., 2015). To quantify the aperiodic components of EEG power spectra, we employed the FOOOF algorithm (*Fitting Oscillations & One-Over-F*)(Donoghue et al., 2020). This method decomposes each channel’s power spectral density (PSD) into an aperiodic component, modeled as a 1/f-like background, and periodic components, corresponding to oscillatory peaks. The aperiodic parameters include the offset, representing broadband power independent of frequency, and the exponent, which characterizes the spectral slope and reflects the balance between neural excitation and inhibition (He, 2014). In our implementation, the function fooofaperiodic() computes the PSD of each EEG channel using the Welch method, with a segment length (nperseg) set to twice the sampling frequency. The PSD is then fitted with the FOOOF model within the 1-30 Hz range, using parameters peak_width_limits=[2,12], max_n_peaks=6, and min_peak_height=0.1. From each fitted model, the aperiodic parameters are extracted using get_params(“aperiodic_params”), providing per-channel estimates of offset and exponent. Channels for which the model did not converge were excluded from subsequent analysis. For statistical comparison across preprocessing conditions (e.g., artifacted data against ICA-based detrending, non-windowed and windowed detrending), Mann-Whitney U tests were applied to the distributions of offset and exponent values, with Bonferroni correction for multiple comparisons. This procedure allowed us to assess the effect of each artifact-removal method on the spectral composition of the EEG signal, ensuring that physiological aperiodic characteristics as much as possible were preserved.

## 3. Results

### 3.1. Extreme capacitive artifacts arise from specific TMS-EEG hardware pairings

We systematically varied combinations of EEG caps (Easycap vs EBneuro cap), amplifiers (BrainAmp vs EBNeuro), and stimulators (EBNeuro vs Nexstim) to identify hardware configurations most prone to generating capacitive artifacts at the stimulation site. Real-time monitoring revealed that changing the stimulator or amplifier alone (datasets D1-D4) did not introduce offset artifacts. In contrast, dataset D7, which used a different EEG cap (EBNeuroCap), showed the pronounced capacitive artifact initially sought. Following acquisition, we pre-processed each dataset as described in the Methods, and computed average TEPs (Figure 2A). We extracted the maximum amplitude per channel to quantify artifact severity (Figure 2B).

To classify the severity of capacitive artifacts, we defined a four-stage outlier system based on Z-score thresholds of channel amplitudes (Lyons, 2013): stage 1 for deviations within 3 sigma, stage 2 for deviations between 3 and 6 sigma, and stage 3 for deviations between 6 and 9 sigma, stage 4 for deviations beyond 9 sigma. Datasets D1-D4 showed channel amplitudes within Stage 1 (≤3 sigma), indicating minimal or no artifact contamination. We classified D5 as Stage 2, with widespread moderate amplitude increase across many channels. We assigned D6 to Stage 3, showing fewer affected channels but with higher peak amplitudes, and D7 to Stage 4, exhibiting severe, localized distortions with extremely high amplitudes and prolonged signal drift. We observed an inverse relationship between the number of affected channels and the magnitude of the artifacts: higher artifact amplitude corresponded to fewer involved channels. This suggests that extreme capacitive artifacts are highly focal, driven by specific stimulator- site interactions, rather than global electrode polarization. The source of the artifact appears to be primarily magnetic perturbation rather than by widespread impedance or electrode contact issues.

### 3.2. Windowed detrending outperforms other methods in managing severe artifacts

Based on the artifact staging, we focused our analysis on the severely contaminated datasets D5-D7 (Figure 1, C-N panels), evaluating three correction strategies: ICA, non-windowed bi-exponential detrending, windowed detrending using polynomial fitting (see Methods).

In dataset D5 (panel C), classified as Stage 2, the raw signal showed moderate capacitive contamination with some residual oscillatory components. ICA-based correction (panel D) effectively removed the early artifact, preserving overall morphology. Non-windowed detrending (panel E) partially recovered the multipeak structure, improving early-phase interpretation. Windowed detrending (panel F) produced comparable results to the non-windowed approach, preserving morphology with better amplitude integrity. In dataset D6 (panel G), corresponding to Stage 3, the artifact was stronger, with slower decay and increased offset. ICA (panel H) suppressed the capacitive artifact restoring physiological components. Non-windowed detrending (panel I) removed the offset but failed to reconstruct the expected initial rising phase, resulting in a downward-trending signal inconsistent with typical TEP morphology. In contrast, windowed detrending (panel J) successfully suppressed the artifact and recovered a recognizable early-phase response. In dataset D7 (panel K), categorized as Stage 4, the raw signal displayed a pronounced downward drift across the entire epoch, resembling the extreme case simulated in Fig. 1C. The ICA-based detrend (panel L) overcorrected the signal, suppressing both artifact and physiological components and leading to a flattened evoked response. Non-windowed detrending (panel M) misfit the signal, introducing artificial zero-centered oscillations around the stimulus onset. Windowed detrending (panel N) yielded the best correction, substantially reducing the drift and partially recovering both early and late oscillatory components, despite some amplitude loss. Notably, all detrending results shown (Figures 2D-N) were produced without ICA. ICA could be applied after detrending to further isolate highlighting informative components. This hybrid approach is expected to enhance TEP quality beyond what ICA or detrending alone can achieve. Overall, windowed detrending demonstrates superior robustness and flexibility in correcting artifacts, particularly in severely contaminated recordings. Its temporally localized fitting allowed accurate capture of non-linear rise and decay profiles, essential for recovering true neurophysiological morphology.

Similar analysis but for the other datasets D1/D2/D3/D4 refer to Supporting Figure 9 and for their ICA detrend components refer to Supporting Figure 10, 11, 12, 13. The amplitude and temporal range in panels was selected as a compromise between interpretability of the signal and value ranges found in the literature. The top of the figure includes a legend describing the hardware configuration for datasets D1-D7.

### 3.3. Windowed detrending as an effective signal denoising strategy

To assess the effect of the different detrending approaches, we estimated the parameters of the aperiodic component, as detrending acts on multiple sources of noise (e.g., the capacitive artifact and other forms) that can overlap with or modulate the neural signal. A good detrending procedure should bring the aperiodic component of the spectrum closer to a physiological 1/f regime with low exponent in log-log coordinates (He, 2014). Here, we compared the changes in offset and slope of the aperiodic components (see Methods 2.4).

For subject D5, the artifacted data (Fig. 3A) showed a mean aperiodic exponent of χ = 1.133 and an offset of −10.96 across 63 channels. ICA-based detrending (Fig. 3B) induced negligible parameter changes (χ = 1.072, offset = −11.12; Uχ = 2131, p = 0.4762, p_adj = 1; Uoffset = 2238, p = 0.217, p_adj = 0.6511). A significant modulation emerged after non-windowed detrending (Fig. 3C), with χ = 0.9254 and offset = −11.27 (Uχ = 2468, p = 0.0184, p_adj = 0.0553; Uoffset = 2832, p = 3.59×10^−5^, p_adj = 1.08×10^−4^). The strongest effect was observed following windowed detrending (Fig. 3D), which reduced χ to 0.3592 and offset to −12.03 (Uχ = 3547, p = 2.51×10^−14^, p_adj = 7.53×10^−14^; Uoffset = 3756, p = 5.57×10^−18^, p_adj = 1.67×10^−17^).

**Figure 3.**
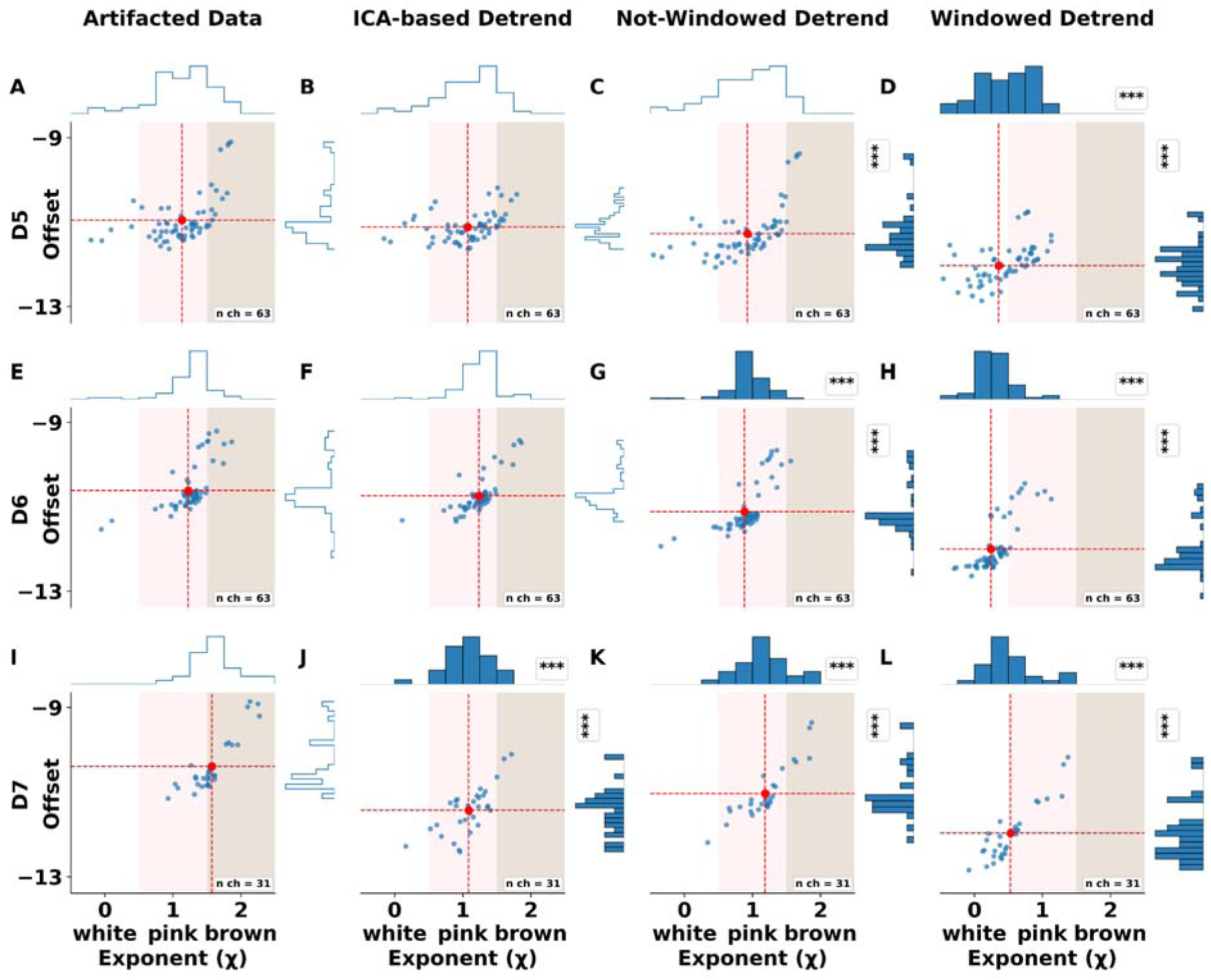
Aperiodic FOOOF parameters across detrending methods. Scatterplots show the relationship between the aperiodic exponent (χ, x-axis) and the offset (y-axis) estimated with the FOOOF algorithm for each subject (rows D5–D7) and preprocessing condition (columns). Each dot corresponds to one EEG channel. Red dashed lines mark the centroid (mean exponent χ and mean offset) of the distribution. Background shading indicates canonical spectral noise regimes: white (χ ≈ 0), pink (χ ≈ 1), and brown (χ ≈ 2). Marginal histograms display the distributions of χ (top) and offset (right). Statistical significance of Mann–Whitney U tests comparing each condition against the artifacted data is shown with asterisks (* p < 0.05; ** p < 0.01; *** p < 0.001). Detrending based on windowed or exponential models systematically reduces the offset and decreases χ toward values compatible with physiological 1/f-like scaling in the post-artifact signal.

For subject D6, the artifacted condition (Fig. 3E) exhibited χ = 1.226 and offset = −10.62 (n = 63). ICA-based detrending (Fig. 3F) again produced no significant changes (χ = 1.235, offset = −10.74; Uχ = 2260, p = 0.1797, p_adj = 0.539; Uoffset = 2384, p = 0.0516, p_adj = 0.155). Both the non-windowed (Fig. 3G) and windowed detrending (Fig. 3H) significantly altered the aperiodic parameters. The former yielded χ = 0.882 and offset = −11.11 (Uχ = 3330, p = 5.29×10^−11^, p_adj = 1.59×10^−10^; Uoffset = 3169, p = 7.61×10^−9^, p_adj = 2.28×10^−8^), while the latter produced χ = 0.2434 and offset = −12.00 (Uχ = 3785, p = 1.60×10^−18^, p_adj = 4.79×10^−18^; Uoffset = 3656, p = 3.55×10^−16^, p_adj = 1.06×10^−15^).

In subject D7, the artifacted condition (Fig. 3I) presented χ = 1.574 and offset = −10.40 (n = 31). ICA-based detrending (Fig. 3J) markedly steepened the aperiodic slope and reduced broadband power (χ = 1.087, offset = −11.44; Uχ = 840, p = 4.32×10^−7^, p_adj = 1.30×10^−6^; Uoffset = 893, p = 6.62×10^−9^, p_adj = 1.99×10^−8^). However, visual inspection revealed that the ICA-filtered spectrum lost its physiological 1/f-like structure, showing excessive suppression of relevant temporal components (Fig. 2L). The non-windowed detrend (Fig. 3K; χ = 1.187, offset = −11.04; Uχ = 774, p = 3.71×10^−5^, p_adj = 1.11×10^−4^; Uoffset = 787, p = 1.65×10^−5^, p_adj = 4.94×10^−5^) partially restored spectral consistency, whereas the windowed detrend (Fig. 3L) achieved both noise attenuation and preservation of the physiological 1/f profile (χ = 0.5343, offset = −11.98; Uχ = 942, p = 8.57×10^−11^, p_adj = 2.57×10^−10^; Uoffset = 914, p = 1.09×10^−9^, p_adj = 3.26×10^−9^).

Globally, the windowed detrending consistently reduced both exponent and offset across all subjects (Δχ ≈ −0.7 to −1.2; Δoffset ≈ −1.0 to −1.5) with high statistical significance (all adjusted p < 10^−6^), while preserving the intrinsic aperiodic structure and physiologically meaningful variability of the EEG spectra.

## 4. Discussion

In this work, we investigated the correction of capacitive artifacts in TMS-EEG using both windowed and non-windowed detrending techniques. Our results indicate that windowed detrending, which separates the rise and decay phases of the capacitive effect, corresponding to electrode charging and discharging, is the most effective approach for managing extreme cases of capacitive artifacts. Among the tested models, polynomial functions outperformed exponential models in both robustness and fitting accuracy.

Previous pipelines have proposed various procedures for handling capacitive artifacts. In the ARTIST pipeline (Wu et al., 2018), detrending is performed prior to epoching, ICA is used to identify and remove components that are mainly contributed by capacitive artifacts, with further rounds of ICA used to eliminate residual noise components. However, algebraic decomposition methods like ICA and principal component analysis (PCA) can become ineffective in the presence of extreme artifacts, which may dominate component space and mask other noise types (Djuwari et al., 2005; Jung et al., 1997, 1998). The TESA pipeline (Mutanen et al., 2020; Rogasch et al., 2017), on the other hand, addresses electrical artifacts, including decay artifacts, recommending their mitigation either via TMS-compatible hardware for online control, or offline methods such as detrending, empirical mode decomposition (EMD), ICA, and PCA. In the case of detrending, TESA uses linear or (bi-) exponential functions, where parameters are estimated from averaged signal per channel and subtracted from each trial. Overall, the use of linear or nonlinear detrending functions can be advantageous under moderate artifact conditions; they often break down when confronted with high-amplitude, nonlinear capacitive trends due to instability in parameter estimation. In contrast, our windowed approach introduces a novel strategy: modeling the rise and decay separately to better reflect the temporal bi-phasic charging-discharging dynamics of capacitive artifacts. This separation enhances convergence during fitting and improves the morphological recovery of TEPs, particularly in the early components most affected by the artifact.

Our results showed that polynomial detrending consistently outperformed exponential fitting. A single exponential function lacks the flexibility required to capture the nonlinear dynamics typical of capacitive artifacts. Accurately modeling these dynamics with exponential functions requires combining multiple exponentials with distinct time constants (e.g., bi- or tri-exponential models), which increases model complexity, reduces interpretability, and can lead to numerical instability (Transtrum et al., 2010). In contrast, low-to-moderate-order polynomial functions inherently offer greater adaptability, as their shape can vary smoothly and non-uniformly across time, making them particularly well-suited for capturing temporally asymmetric signal features. That said, very high-order polynomials can still lead to overfitting, especially in the decay phase, potentially distorting or removing meaningful oscillatory activity (Kim et al., 2018; Laishram et al., 2024). Although multi-component exponential models may match the performance of polynomials in some cases, they do so at the expense of increased computational and analytical burden. Within the moderate polynomial orders used in this study (3rd order), the fitting remained stable and effective across a wide range of artifact profiles. In fact, the decay phase should be fitted with a low-degree polynomial, as higher-order fitting may lead to overfitting, potentially removing oscillatory components or incorporating contaminated data. The goal of detrending is to eliminate the noisy trend that distorts the TEP dynamics, not to alter the underlying signal. In contrast, the rise phase, being fast and impulsive, strongly contaminates the first milliseconds after stimulation, making it extremely difficult to preserve the true neural response to TMS. Therefore, careful control of polynomial order is critical, tailoring it to the artifact phase: high-order for rise, low-order for decay.

The analysis of the aperiodic component further supported the windowed detrending as an effective denoising strategy compared to non-windowed and ICA-based corrections. TMS-EEG traces affected by artifacts exhibited inflated offsets and steep aperiodic exponents (Figure 3), consistent with broadband spectral contamination from residual capacitive decay. In contrast, windowed detrending produced the lowest aperiodic parameters, which were at the boundary of white noise and pink-noise regime, as should be generated by brain networks (Allegrini et al., 2009; Bédard et al., 2006; West, 2010). Nevertheless, caution is warranted: although moderate-order polynomials (second- to third-order) provide sufficient flexibility to capture baseline curvature, higher-order fits may overcompensate for slow trends, potentially attenuating or distorting genuine low-frequency and oscillatory components. Thus, the improved aperiodic stability obtained with windowed detrending highlights both its efficacy and the importance of controlling polynomial order to avoid excessive spectral flattening.

In this work, we treated capacitive artifacts as a distinct subclass of stimulation-related electrical artifacts. Although the precise nature of these artifacts remains debated (Ilmoniemi & Kičić, 2010; Litvak et al., 2007), several biophysical models have been proposed to explain their origin, often implicating the passive tissue properties of the scalp, skull, and electrode interfaces (Freche et al., 2018). These models suggest that nonlinear mitigation strategies, particularly those based on power-law dynamics, may better capture the temporal profile of capacitive artifacts. While beyond the scope of the present study, such approaches may inform biophysically grounded detrending strategies.

Our method, like most model-based approaches, is data-driven, relying on curve-fitting to estimated model parameters for each trial. However, this introduces potential computational challenges, especially in large datasets or complex models. A promising extension of this methodology is the adoption of generative models able to determine in advance a set of candidate templates for the capacitive artifact. In this latter case, the parameter estimation step is replaced by a shape-matching procedure between the morphology of individual trials and a catalog of prototypical artifacts (see Supporting Figure 1 for an example of capacitive artifact generation). In this framework, artifact subtraction is achieved by shape matching, rather than parametric fitting. This opens the door to machine learning-assisted classification, where neural networks or clustering algorithms select the optimal artifact template (Buccino et al., 2018; Griffiths et al., 2022; Keihani et al., 2025; Nielsen et al., 2020; Park & Kim, 2022; Srichawla, 2024; Williams, 2024). While a template-based approach may improve the quality of artifact correction, it comes at the cwost of increased computational load, due to the need to generate a sufficiently comprehensive set of synthetic artifacts.

These prototypes must span the multidimensional parameter space of the underlying capacitive model, including any constraints, thus introducing combinatorial complexity.

We also explored hardware-induced variability in capacitive artifact severity. This analysis revealed that the generation of capacitive artifacts is not only a function of EEG sensor, but also depends on electrode mounting configuration, amplifiers, mechanical stability, and electrical shielding. Datasets with severe artifacts typically showed localized effects, strongly affecting only a few channels near the stimulation site. These artifacts obscure early TEP components, which are crucial for tracking cortical excitability and network-level propagation (Momi et al., 2023, 2025). Therefore, differences in electrode design or in the physical coupling between electrode and scalp (Huigen et al., 2002; Onaral et al., 1984; R. Eggins, 1993; Tallgren et al., 2005; Varone et al., 2021; Veniero et al., 2009; Vernet & Thut, 2014; J. Virtanen et al., 1999) likely play a central role in shaping the amplitude and persistence of the observed capacitive artifacts.

## 5. Conclusion

Our results reinforce the notion that the employment of optimal, TMS-compatible hardware and careful online prevention and monitoring of capacitive artifacts remain the most effective strategy to ensure high-quality TMS-EEG data. Nevertheless, when optimal acquisition conditions cannot be guaranteed, the use of reliable offline correction algorithms becomes essential and should be considered an integral component of the processing framework.

Different analysis pipelines can yield divergent results, highlighting the importance of robust and well-defined preprocessing strategies for correcting stimulation-related artifacts in TMS-EEG. We showed that windowed detrending, especially with polynomial fitting, offers a reliable and flexible solution for correcting extreme capacitive artifacts, outperforming ICA and non-windowed models in signal reconstruction and morphology preservation across different settings. These findings reinforce the notion that preprocessing decisions significantly shape the resulting TEPs, especially in pipelines known to exhibit variability (Belardinelli et al., 2019; Bertazzoli et al., 2021; Brancaccio et al., 2024; Casarotto et al., 2010; Kuhn et al., 2025; Lioumis et al., 2009; Siebner et al., 2019; Vucic et al., 2023). As such, adopting standardized, validated artifact correction methods is vital for improving test–retest reliability and cross-study comparability (Bertazzoli et al., 2025).

Our suggestion is that, if the ICA decomposition appears dominated by extreme artifacts, thereby becoming insensitive or unintelligible to other physiological or non-physiological components, we recommend applying windowed detrending prior to ICA. This preprocessing step can effectively reduce large-scale drifts and non-stationarities, stabilizing the decomposition and improving the separation of meaningful components.

The implications are particularly relevant for clinical applications of TMS-EEG, where both reproducibility and effective artifact suppression are essential for extracting reliable neural biomarkers. Refined preprocessing pipelines are especially valuable in the context of neurological and neuropsychiatric disorders (Bae et al., 2007; Baumer et al., 2020; Gefferie et al., 2023; Lorentzen et al., 2022; She et al., 2024; Watanabe & Yamasue, 2025) where subtle alterations in cortical excitability or connectivity may otherwise be masked or misinterpreted due to residual stimulation-related artifacts. Ultimately, enhancing the accuracy and interpretability of TMS-EEG data will be key to strengthening its translational potential in diagnosis, patient stratification, treatment monitoring across a range of clinical populations and basic understanding of TMS-based perturbative approaches (Casula et al., 2017; Couto et al., 2025; D’Ambrosio et al., 2022; Fecchio et al., 2025; Russo et al., 2025; Vallesi et al., 2021).

## Data statement

All the data are available upon request and the code is available at GitHub repository link https://github.com/albertoarturovergani/windowedDetrend.

## Data Availability

All the data are available upon request.

## Acknowledgements

We thank Gabriel Hassan, Elisabetta Litterio, and Giulia Furregoni for their contribution to data acquisition in Milan.

## References

Allegrini, P., Menicucci, D., Bedini, R., Fronzoni, L., Gemignani, A., Grigolini, P., West, B. J., & Paradisi, P. (2009). Spontaneous brain activity as a source of ideal 1 / f noise. Physical Review E, 80(6), 061914. 10.1103/PhysRevE.80.061914

Atti, I., Belardinelli, P., Ilmoniemi, R. J., & Metsomaa, J. (2024). Measuring the accuracy of ICA-based artifact removal from TMS-evoked potentials. Brain Stimulation, 17(1), 10–18. 10.1016/j.brs.2023.12.001

Bae, E. H., Schrader, L. M., Machii, K., Alonso-Alonso, M., Riviello, J. J., Pascual-Leone, A., & Rotenberg, A. (2007). Safety and tolerability of repetitive transcranial magnetic stimulation in patients with epilepsy: A review of the literature. Epilepsy & Behavior, 10(4), 521–528. 10.1016/j.yebeh.2007.03.004

Baumer, F. M., Pfeifer, K., Fogarty, A., Pena-Solorzano, D., Rolle, C. E., Wallace, J. L., Rotenberg, A., & Fisher, R. S. (2020). Cortical Excitability, Synaptic Plasticity, and Cognition in Benign Epilepsy With Centrotemporal Spikes: A Pilot TMS-EMG-EEG Study. Journal of Clinical Neurophysiology, 37(2), 170. 10.1097/WNP.0000000000000662

Bédard, C., Kröger, H., & Destexhe, A. (2006). Does the $1/f$ Frequency Scaling of Brain Signals Reflect Self-Organized Critical States? Physical Review Letters, 97(11), 118102. 10.1103/PhysRevLett.97.118102

Belardinelli, P., Biabani, M., Blumberger, D. M., Bortoletto, M., Casarotto, S., David, O., Desideri, D., Etkin, A., Ferrarelli, F., Fitzgerald, P. B., Fornito, A., Gordon, P. C., Gosseries, O., Harquel, S., Julkunen, P., Keller, C. J., Kimiskidis, V. K., Lioumis, P., Miniussi, C., … Ilmoniemi, R. J. (2019). Reproducibility in TMS–EEG studies: A call for data sharing, standard procedures and effective experimental control. Brain Stimulation, 12(3), 787–790. 10.1016/j.brs.2019.01.010

Bertazzoli, G., Dognini, E., Fried, P. J., Miniussi, C., Julkunen, P., & Bortoletto, M. (2025). Bridging the gap to clinical use: A systematic review on TMS–EEG test-retest reliability. Clinical Neurophysiology, 171, 133–145. 10.1016/j.clinph.2025.01.002

Bertazzoli, G., Esposito, R., Mutanen, T. P., Ferrari, C., Ilmoniemi, R. J., Miniussi, C., & Bortoletto, M. (2021). The impact of artifact removal approaches on TMS-EEG signal. NeuroImage, 239, 118272. 10.1016/j.neuroimage.2021.118272

Brancaccio, A., Tabarelli, D., Zazio, A., Bertazzoli, G., Metsomaa, J., Ziemann, U., Bortoletto, M., & Belardinelli, P. (2024). Towards the definition of a standard in TMS-EEG data preprocessing. NeuroImage, 301, 120874. 10.1016/j.neuroimage.2024.120874

Buccino, A. P., Kordovan, M., Ness, T. V., Merkt, B., Häfliger, P. D., Fyhn, M., Cauwenberghs, G., Rotter, S., & Einevoll, G. T. (2018). Combining biophysical modeling and deep learning for multielectrode array neuron localization and classification. Journal of Neurophysiology, 120(3), 1212–1232. 10.1152/jn.00210.2018

Burbank, D. P., & Webster, J. G. (1978). Reducing skin potential motion artefact by skin abrasion. Medical and Biological Engineering and Computing, 16(1), 31–38. 10.1007/BF02442929

Casarotto, S., Fecchio, M., Rosanova, M., Varone, G., D’Ambrosio, S., Sarasso, S., Pigorini, A., Russo, S., Comanducci, A., Ilmoniemi, R. J., & Massimini, M. (2022). The rt-TEP tool: Real-time visualization of TMS-Evoked Potentials to maximize cortical activation and minimize artifacts. Journal of Neuroscience Methods, 370, 109486. 10.1016/j.jneumeth.2022.109486

Casarotto, S., Lauro, L. J. R., Bellina, V., Casali, A. G., Rosanova, M., Pigorini, A., Defendi, S., Mariotti, M., & Massimini, M. (2010). EEG Responses to TMS Are Sensitive to Changes in the Perturbation Parameters and Repeatable over Time. PLOS ONE, 5(4), e10281. 10.1371/journal.pone.0010281

Casula, E. P., Bertoldo, A., Tarantino, V., Maiella, M., Koch, G., Rothwell, J. C., Toffolo, G. M., & Bisiacchi, P. S. (2017). TMS-evoked long-lasting artefacts: A new adaptive algorithm for EEG signal correction. Clinical Neurophysiology: Official Journal of the International Federation of Clinical Neurophysiology, 128(9), 1563– 1574. 10.1016/j.clinph.2017.06.003

Conde, V., Tomasevic, L., Akopian, I., Stanek, K., Saturnino, G. B., Thielscher, A., Bergmann, T. O., & Siebner, H. R. (2019). The non-transcranial TMS-evoked potential is an inherent source of ambiguity in TMS-EEG studies. NeuroImage, 185, 300–312. 10.1016/j.neuroimage.2018.10.052

Couto, B. A., Fecchio, M., Russo, S., Martino, E. D., Parmigiani, S., Sarasso, S., Graven-Nielsen, T., Andrade, D. C. de, Massimini, M., Rosanova, M., & Casali, A. G. (2025). Extracting Reproducible Components from Electroencephalographic Responses to Transcranial Magnetic Stimulation with Group Task-Related Component Analysis (p. 2025.06.02.657489). bioRxiv. 10.1101/2025.06.02.657489

D’Ambrosio, S., Jiménez-Jiménez, D., Silvennoinen, K., Zagaglia, S., Perulli, M., Poole, J., Comolatti, R., Fecchio, M., Sisodiya, S. M., & Balestrini, S. (2022). Physiological symmetry of transcranial magnetic stimulation-evoked EEG spectral features. Human Brain Mapping, 43(18), 5465–5477. 10.1002/hbm.26022

Djuwari, D., Kumar, D. K., & Palaniswami, M. (2005). Limitations of ICA for Artefact Removal. 2005 IEEE Engineering in Medicine and Biology 27th Annual Conference, 4685–4688. 10.1109/IEMBS.2005.1615516

Donoghue, T., Haller, M., Peterson, E. J., Varma, P., Sebastian, P., Gao, R., Noto, T., Lara, A. H., Wallis, J. D., Knight, R. T., Shestyuk, A., & Voytek, B. (2020). Parameterizing neural power spectra into periodic and aperiodic components. Nature Neuroscience, 23(12), 1655–1665. 10.1038/s41593-020-00744-x

Fecchio, M., Russo, S., Couto, B. A., Mikulan, E., Pigorini, A., Furregoni, G., Hassan, G., D’Ambrosio, S., Solbiati, M., Viganò, A., Parmigiani, S., Sarasso, S., Casarotto, S., Massimini, M., Casali, A. G., & Rosanova, M. (2025). The specific spatiotemporal evolution of TMS-evoked potentials reflects the engagement of cortical circuits (p. 2025.06.25.661535). bioRxiv. 10.1101/2025.06.25.661535

Freche, D., Naim-Feil, J., Peled, A., Levit-Binnun, N., & Moses, E. (2018). A quantitative physical model of the TMS- induced discharge artifacts in EEG. PLOS Computational Biology, 14(7), e1006177. 10.1371/journal.pcbi.1006177

Gefferie, S. R., Jiménez-Jiménez, D., Visser, G. H., Helling, R. M., Sander, J. W., Balestrini, S., & Thijs, R. D. (2023). Transcranial magnetic stimulation-evoked electroencephalography responses as biomarkers for epilepsy: A review of study design and outcomes. Human Brain Mapping, 44(8), 3446–3460. 10.1002/hbm.26260

Griffiths, J. D., Wang, Z., Ather, S. H., Momi, D., Rich, S., Diaconescu, A., McIntosh, A. R., & Shen, K. (2022). Deep Learning-Based Parameter Estimation for Neurophysiological Models of Neuroimaging Data. bioRxiv. 10.1101/2022.05.19.492664

He, B. J. (2014). Scale-free brain activity: Past, present, and future. Trends in Cognitive Sciences, 18(9), 480–487. 10.1016/j.tics.2014.04.003

Hernandez-Pavon, J. C., Kugiumtzis, D., Zrenner, C., Kimiskidis, V. K., & Metsomaa, J. (2022). Removing artifacts from TMS-evoked EEG: A methods review and a unifying theoretical framework. Journal of Neuroscience Methods, 376, 109591. 10.1016/j.jneumeth.2022.109591

Huigen, E., Peper, A., & Grimbergen, C. A. (2002). Investigation into the origin of the noise of surface electrodes. Medical and Biological Engineering and Computing, 40(3), 332–338. 10.1007/BF02344216

Ilmoniemi, R. J., Hernandez-Pavon, J. C., Mäkelä, N. N., Metsomaa, J., Mutanen, T. P., Stenroos, M., & Sarvas, J. (2015). Dealing with artifacts in TMS-evoked EEG. 2015 37th Annual International Conference of the IEEE Engineering in Medicine and Biology Society (EMBC), 230–233. 10.1109/EMBC.2015.7318342

Ilmoniemi, R. J., & Kičić, D. (2010). Methodology for Combined TMS and EEG. Brain Topography, 22(4), 233–248. 10.1007/s10548-009-0123-4

Johnson, J. B. (1928). Thermal Agitation of Electricity in Conductors. Physical Review, 32(1), 97–109. 10.1103/PhysRev.32.97

Jung, T.-P., Humphries, C., Lee, T.-W., Makeig, S., McKeown, M., Iragui, V., & Sejnowski, T. J. (1997). Extended ICA Removes Artifacts from Electroencephalographic Recordings. Advances in Neural Information Processing Systems, 10. https://proceedings.neurips.cc/paper/1997/hash/674bfc5f6b72706fb769f5e93667bd23-Abstract.html

Jung, T.-P., Humphries, C., Lee, T.-W., Makeig, S., McKeown, M. J., Iragui, V., & Sejnowski, T. J. (1998). Removing electroencephalographic artifacts: Comparison between ICA and PCA. Neural Networks for Signal Processing VIII. Proceedings of the 1998 IEEE Signal Processing Society Workshop (Cat. No.98TH8378), 63–72. 10.1109/NNSP.1998.710633

Keihani, A., Donati, F. L., Russo, S., Parmigiani, S., Solbiati, M., Casali, A. G., Fecchio, M., Chaichian, O., Rothwell, J., Massimini, M., Rocchi, L., Rosanova, M., & Ferrarelli, F. (2025). Recognizing EEG responses to active TMS vs. sham stimulations in different TMS-EEG datasets: A machine learning approach (p. 2025.06.29.662189). bioRxiv. 10.1101/2025.06.29.662189

Kim, J.-H., Su, W., & Song, Y. J. (2018). On Stability of a Polynomial. Journal of Applied Mathematics & Informatics, 36(3_4), 231–236. 10.14317/jami.2018.231

Kuhn, T., Indahlastari, A., Vila-Rodriguez, F., Fonzo, G., Petersen, N., Rotstein, N., Davis, T., Halavi, S., Mehler, D., Repple, J., Hahn, T., Bertazzoli, G., Reggente, N., Woods, A., Dang, B. H., Mason, X., Laidi, C., Chou, T., Bortoletto, M., … Thompson, P. (2025). The ENIGMA-Neuromodulation working group – A mission statement. Brain Stimulation, 18(2), 142–144. 10.1016/j.brs.2024.12.1477

Laishram, S., Sarma, R., & Sharma, H. (2024). Stability of certain higher degree polynomials. International Journal of Number Theory, 20(01), 229–240. 10.1142/S1793042124500118

Lentka, Ł., & Smulko, J. (2019). Methods of trend removal in electrochemical noise data – Overview. Measurement, 131, 569–581. 10.1016/j.measurement.2018.08.023

Lioumis, P., Kičić, D., Savolainen, P., Mäkelä, J. P., & Kähkönen, S. (2009). Reproducibility of TMS—Evoked EEG responses. Human Brain Mapping, 30(4), 1387–1396. 10.1002/hbm.20608

Litvak, V., Komssi, S., Scherg, M., Hoechstetter, K., Classen, J., Zaaroor, M., Pratt, H., & Kahkonen, S. (2007). Artifact correction and source analysis of early electroencephalographic responses evoked by transcranial magnetic stimulation over primary motor cortex. NeuroImage, 37(1), 56–70. 10.1016/j.neuroimage.2007.05.015

Lorentzen, R., Nguyen, T. D., McGirr, A., Hieronymus, F., & Østergaard, S. D. (2022). The efficacy of transcranial magnetic stimulation (TMS) for negative symptoms in schizophrenia: A systematic review and meta-analysis. Schizophrenia, 8(1), 35. 10.1038/s41537-022-00248-6

Lyons, L. (2013). Discovering the Significance of 5 sigma (No. arXiv:1310.1284). arXiv. 10.48550/arXiv.1310.1284

Ma, J., Tao, P., Bayram, S., & Svetnik, V. (2012). Muscle artifacts in multichannel EEG: Characteristics and reduction. Clinical Neurophysiology, 123(8), 1676–1686. 10.1016/j.clinph.2011.11.083

Miniussi, C., & Thut, G. (2010). Combining TMS and EEG Offers New Prospects in Cognitive Neuroscience. Brain Topography, 22(4), 249–256. 10.1007/s10548-009-0083-8

Momi, D., Wang, Z., & Griffiths, J. D. (2023). TMS-evoked responses are driven by recurrent large-scale network dynamics. eLife, 12, e83232. 10.7554/eLife.83232

Momi, D., Wang, Z., Parmigiani, S., Mikulan, E., Bastiaens, S. P., Oveisi, M. P., Kadak, K., Gaglioti, G., Waters, A. C., Hill, S., Pigorini, A., Keller, C. J., & Griffiths, J. D. (2025). Stimulation mapping and whole-brain modeling reveal gradients of excitability and recurrence in cortical networks. Nature Communications, 16(1), 3222. 10.1038/s41467-025-58187-6

Mutanen, T. P., Biabani, M., Sarvas, J., Ilmoniemi, R. J., & Rogasch, N. C. (2020). Source-based artifact-rejection techniques available in TESA, an open-source TMS-EEG toolbox. Brain Stimulation, 13(5), 1349–1351. 10.1016/j.brs.2020.06.079

Mutanen, T. P., Kukkonen, M., Nieminen, J. O., Stenroos, M., Sarvas, J., & Ilmoniemi, R. J. (2016). Recovering TMS- evoked EEG responses masked by muscle artifacts. NeuroImage, 139, 157–166. 10.1016/j.neuroimage.2016.05.028

Mutanen, T. P., Metsomaa, J., Makkonen, M., Varone, G., Marzetti, L., & Ilmoniemi, R. J. (2022). Source-based artifact- rejection techniques for TMS-EEG. Journal of Neuroscience Methods, 382, 109693. 10.1016/j.jneumeth.2022.109693

Nielsen, A. N., Barch, D. M., Petersen, S. E., Schlaggar, B. L., & Greene, D. J. (2020). Machine Learning With Neuroimaging: Evaluating Its Applications in Psychiatry. Biological Psychiatry: Cognitive Neuroscience and Neuroimaging, 5(8), 791–798. 10.1016/j.bpsc.2019.11.007

Nyquist, H. (1928). Thermal Agitation of Electric Charge in Conductors. Physical Review, 32(1), 110–113. 10.1103/PhysRev.32.110

Onaral, B., Sun, H. H., & Schwan, H. P. (1984). Electrical Properties of Bioelectrodes. IEEE Transactions on Biomedical Engineering, BME-31(12), 827–832. 10.1109/TBME.1984.325245

Park, D., & Kim, I. (2022). Application of Machine Learning in the Field of Intraoperative Neurophysiological Monitoring: A Narrative Review. Applied Sciences, 12(15), Articolo 15. 10.3390/app12157943

R. Eggins, B. (1993). Skin contact electrodes for medical applications. Analyst, 118(4), 439–442. 10.1039/AN9931800439

Rogasch, N. C., Sullivan, C., Thomson, R. H., Rose, N. S., Bailey, N. W., Fitzgerald, P. B., Farzan, F., & Hernandez-Pavon, J. C. (2017). Analysing concurrent transcranial magnetic stimulation and electroencephalographic data: A review and introduction to the open-source TESA software. NeuroImage, 147, 934–951. 10.1016/j.neuroimage.2016.10.031

Ruddy, K. L., Woolley, D. G., Mantini, D., Balsters, J. H., Enz, N., & Wenderoth, N. (2018). Improving the quality of combined EEG-TMS neural recordings: Introducing the coil spacer. Journal of Neuroscience Methods, 294, 34–39. 10.1016/j.jneumeth.2017.11.001

Russo, S., Claar, L. D., Furregoni, G., Marks, L. C., Krishnan, G., Zauli, F. M., Hassan, G., Solbiati, M., d’Orio, P., Mikulan, E., Sarasso, S., Rosanova, M., Sartori, I., Bazhenov, M., Pigorini, A., Massimini, M., Koch, C., & Rembado, I. (2025). Thalamic feedback shapes brain responses evoked by cortical stimulation in mice and humans. Nature Communications, 16(1), 3627. 10.1038/s41467-025-58717-2

Serway, R. A., & Jewett, J. W. (con un contributo di Internet Archive). (2004). Physics for scientists and engineers. Belmont, CA : Thomson-Brooks/Cole. http://archive.org/details/physicssciengv2p00serw

She, X., Nix, K. C., Cline, C. C., Qi, W., Tugin, S., He, Z., & Baumer, F. M. (2024). Stability of transcranial magnetic stimulation electroencephalogram evoked potentials in pediatric epilepsy. Scientific Reports, 14(1), 9045. 10.1038/s41598-024-59468-8

Siebner, H. R., Conde, V., Tomasevic, L., Thielscher, A., & Bergmann, T. O. (2019). Distilling the essence of TMS-evoked EEG potentials (TEPs): A call for securing mechanistic specificity and experimental rigor. Brain Stimulation, 12(4), 1051–1054. 10.1016/j.brs.2019.03.076

Siebner, H. R., Funke, K., Aberra, A. S., Antal, A., Bestmann, S., Chen, R., Classen, J., Davare, M., Di Lazzaro, V., Fox, P. T., Hallett, M., Karabanov, A. N., Kesselheim, J., Beck, M. M., Koch, G., Liebetanz, D., Meunier, S., Miniussi, C., Paulus, W., … Ugawa, Y. (2022). Transcranial magnetic stimulation of the brain: What is stimulated? – A consensus and critical position paper. Clinical Neurophysiology, 140, 59–97. 10.1016/j.clinph.2022.04.022

Srichawla, B. S. (2024). Future of neurocritical care: Integrating neurophysics, multimodal monitoring, and machine learning. World Journal of Critical Care Medicine, 13(2), 91397. 10.5492/wjccm.v13.i2.91397

Stango, A., Zazio, A., Barchiesi, G., Bonfiglio, N. S., & Bortoletto, M. (2025). High-frequency sampling rate reduces TMS- pulse artifact duration but not decay artifact: Implications for immediate TMS-EEG responses (p. 2025.03.05.641655). bioRxiv. 10.1101/2025.03.05.641655

Tallgren, P., Vanhatalo, S., Kaila, K., & Voipio, J. (2005). Evaluation of commercially available electrodes and gels for recording of slow EEG potentials. Clinical Neurophysiology, 116(4), 799–806. 10.1016/j.clinph.2004.10.001

Transtrum, M. K., Machta, B. B., & Sethna, J. P. (2010). Why are Nonlinear Fits to Data so Challenging? Physical Review Letters, 104(6), 060201. 10.1103/PhysRevLett.104.060201

Vafidis, P., Kimiskidis, V. K., & Kugiumtzis, D. (2019). Evaluation of algorithms for correction of transcranial magnetic stimulation-induced artifacts in electroencephalograms. Medical & Biological Engineering & Computing, 57(12), 2599–2615. 10.1007/s11517-019-02053-3

Vallesi, A., Del Felice, A., Capizzi, M., Tafuro, A., Formaggio, E., Bisiacchi, P., Masiero, S., & Ambrosini, E. (2021). Natural oscillation frequencies in the two lateral prefrontal cortices induced by Transcranial Magnetic Stimulation. NeuroImage, 227, 117655. 10.1016/j.neuroimage.2020.117655

Varone, G., Hussain, Z., Sheikh, Z., Howard, A., Boulila, W., Mahmud, M., Howard, N., Morabito, F. C., & Hussain, A. (2021). Real-Time Artifacts Reduction during TMS-EEG Co-Registration: A Comprehensive Review on Technologies and Procedures. Sensors, 21(2), Articolo 2. 10.3390/s21020637

Veniero, D., Bortoletto, M., & Miniussi, C. (2009). TMS-EEG co-registration: On TMS-induced artifact. Clinical Neurophysiology, 120(7), 1392–1399. 10.1016/j.clinph.2009.04.023

Vernet, M., & Thut, G. (2014). Electroencephalography During Transcranial Magnetic Stimulation: Current Modus Operandi. In A. Rotenberg, J. C. Horvath, & A. Pascual-Leone (A c. Di), Transcranial Magnetic Stimulation (pp. 197–232). Springer. 10.1007/978-1-4939-0879-0_11

Virtanen, J., Ruohonen, J., Näätänen, R., & Ilmoniemi, R. J. (1999). Instrumentation for the measurement of electric brain responses to transcranial magnetic stimulation. Medical & Biological Engineering & Computing, 37(3), 322–326. 10.1007/BF02513307

Virtanen, P., Gommers, R., Oliphant, T. E., Haberland, M., Reddy, T., Cournapeau, D., Burovski, E., Peterson, P., Weckesser, W., Bright, J., van der Walt, S. J., Brett, M., Wilson, J., Millman, K. J., Mayorov, N., Nelson, A. R. J., Jones, E., Kern, R., Larson, E., … van Mulbregt, P. (2020). SciPy 1.0: Fundamental algorithms for scientific computing in Python. Nature Methods, 17(3), 261–272. 10.1038/s41592-019-0686-2

Voytek, B., Kramer, M. A., Case, J., Lepage, K. Q., Tempesta, Z. R., Knight, R. T., & Gazzaley, A. (2015). Age-Related Changes in 1/f Neural Electrophysiological Noise. Journal of Neuroscience, 35(38), 13257–13265. 10.1523/JNEUROSCI.2332-14.2015

Vucic, S., Stanley Chen, K.-H., Kiernan, M. C., Hallett, M., Benninger, David. H., Di Lazzaro, V., Rossini, P. M., Benussi, A., Berardelli, A., Currà, A., Krieg, S. M., Lefaucheur, J.-P., Long Lo, Y., Macdonell, R. A., Massimini, M., Rosanova, M., Picht, T., Stinear, C. M., Paulus, W., … Chen, R. (2023). Clinical diagnostic utility of transcranial magnetic stimulation in neurological disorders. Updated report of an IFCN committee. Clinical Neurophysiology, 150, 131–175. 10.1016/j.clinph.2023.03.010

Watanabe, T., & Yamasue, H. (2025). Noninvasive reduction of neural rigidity alters autistic behaviors in humans. Nature Neuroscience, 28(6), 1348–1360. 10.1038/s41593-025-01961-y

West, B. J. (2010). Fractal Physiology and the Fractional Calculus: A Perspective. Frontiers in Physiology, 1. 10.3389/fphys.2010.00012

Williams, K. S. (2024). Evaluations of artificial intelligence and machine learning algorithms in neurodiagnostics. Journal of Neurophysiology, 131(5), 825–831. 10.1152/jn.00404.2023

Wu, W., Keller, C. J., Rogasch, N. C., Longwell, P., Shpigel, E., Rolle, C. E., & Etkin, A. (2018a). ARTIST: A fully automated artifact rejection algorithm for single-pulse TMS-EEG data. Human Brain Mapping, 39(4), 1607–1625. 10.1002/hbm.23938

Xu, N., Shang, P., & Kamae, S. (2009). Minimizing the effect of exponential trends in detrended fluctuation analysis. Chaos, Solitons & Fractals, 41(1), 311–316. 10.1016/j.chaos.2007.12.006

