## Supporting Materials for "Effective correction of extreme capacitive artifacts in TMS-EEG via windowed detrending"

### Supporting material

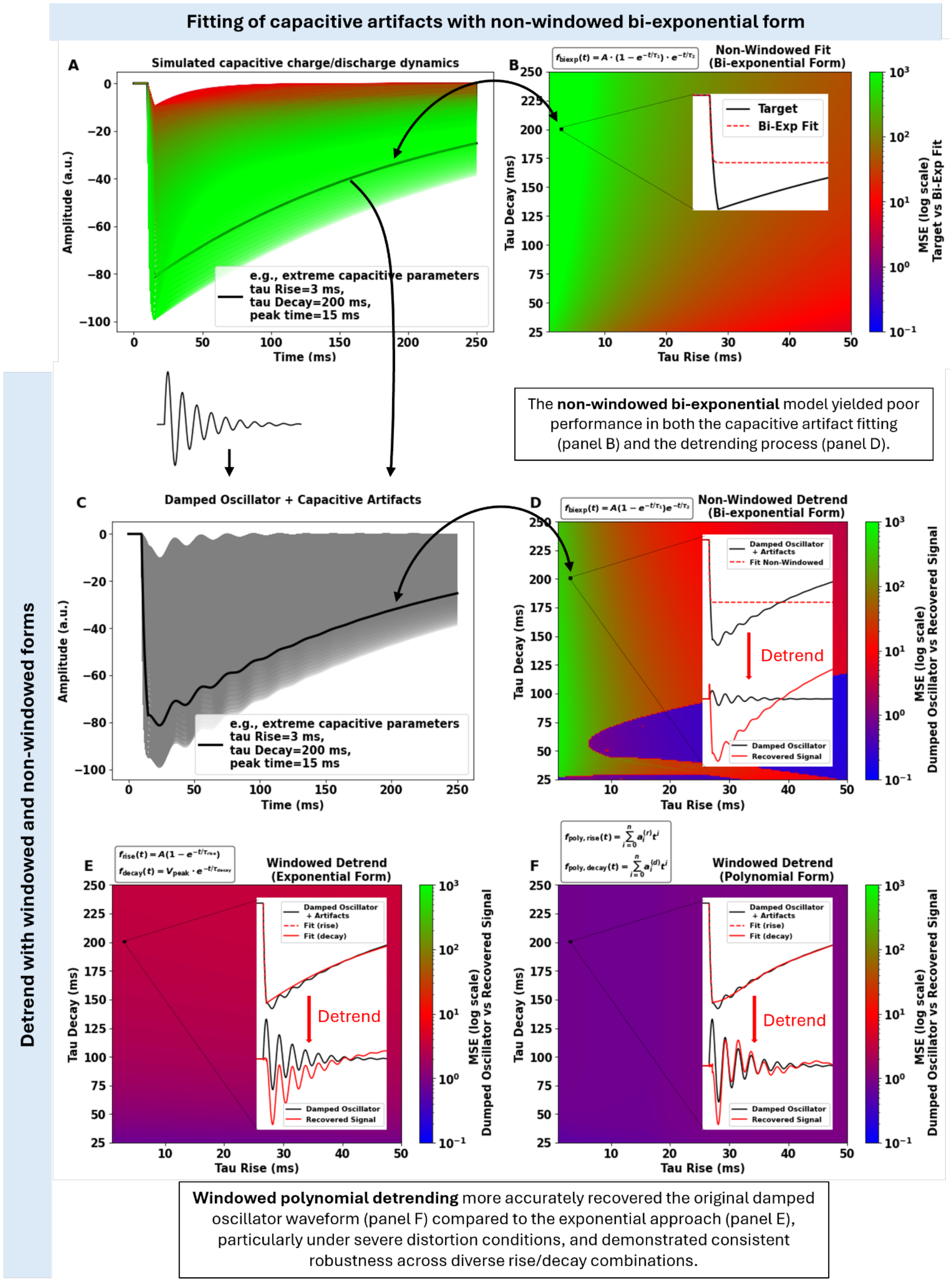

**Supporting Figure 1. Technical validation of windowed detrending across a broad parameter space of capacitive artifacts.** **(A)** Simulated capacitive artifacts modeled as exponential rise and decay functions, with independent variation of the time constants. The extreme artifact case used in Figure 1 is highlighted (tau rise = 3 ms, tau decay = 200 ms, peak latency= 15 ms). **(B)** Mean squared error (MSE, log scale) from fitting each artifact in (A) using a non-windowed bi-exponential model applied directly to capacitive artifacts across the parameter space. Poor performance is observed in the extreme artifact region. Inset: example of poor fitting (target in black and fitted curve in red dashed). **(C)** Contaminated signals generated by adding each artifact from (A) to a simulated damped oscillator. **(D)** Detrending performance using non-windowed bi-exponential models, applied to contaminated signals from (C). Poor signal recovery is observed, especially for extreme artifacts (inset). **(E)** Windowed exponential detrending, applied separately to rise and decay phases, improves model convergence and reduces fitting errors across the parameter space. **(F)** Windowed polynomial detrending shows superior performance, especially under extreme artifact conditions, further enhancing the recovery of the original neural signal.

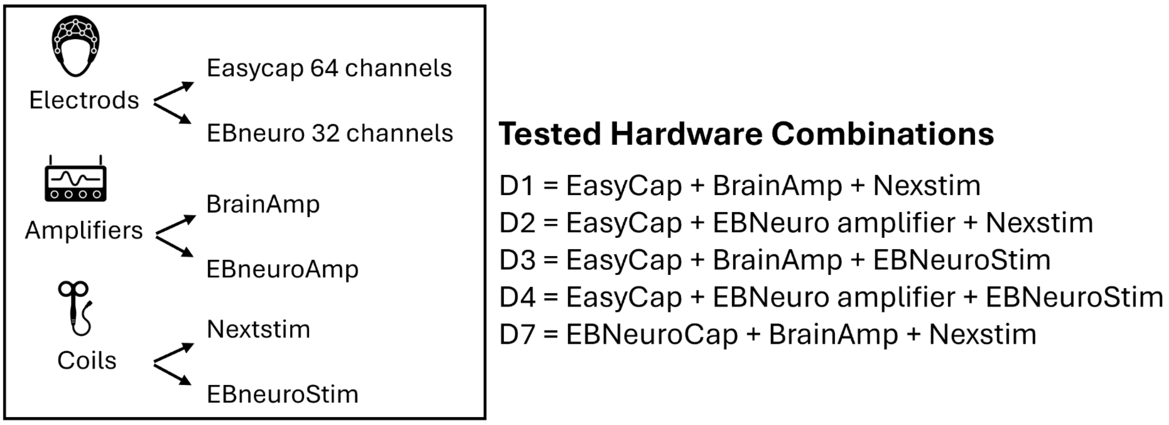


**Supporting Figure 2. Overview of hardware combinations obtained by varying electrodes, amplifiers, and coils, with the aim of isolating the source of capacitive artifacts**. Datasets D5 and D6 are not included in this figure, as they were collected at the Chalfont Centre for Epilepsy (Buckinghamshire, UK) from already published work (D'Ambrosio et al., 2022). The numbering from D1 to D7 reflects the relative importance of the capacitive artifact rather than the order of acquisition (see Figure 2A/B).

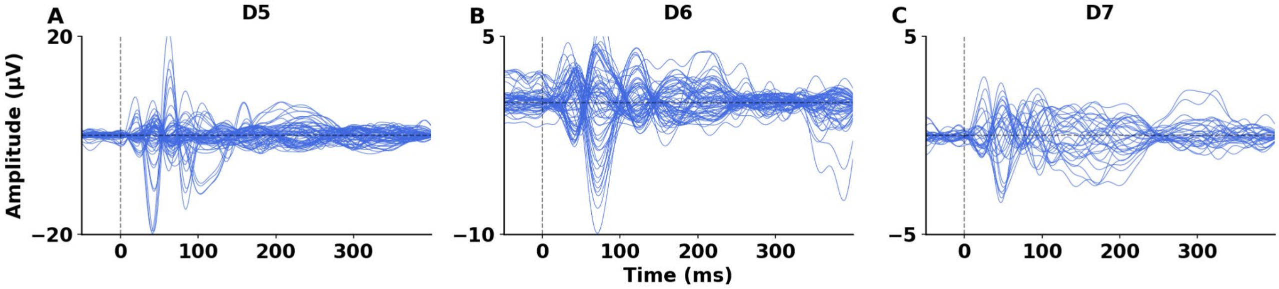


**Supporting Figure 3. Windowed exponential detrending applied to real datasets with increasing levels of capacitive artifact severity.** Panels A–C show detrended EEG traces using windowed exponential fitting for dataset D5 (stage 2), D6 (stage 3), and D7 (stage 4). Signals are shown in microvolts (µV) over time (ms), across channels. The raw, artifact-contaminated signals corresponding to these datasets are shown in Figure 2 (panel C, G, K). Each artifact was modeled using separate exponential functions for the rise and decay phases within temporal windows defined using the procedure described in the Methods. This approach effectively attenuated sharp transients while preserving the physiological TEP structure, particularly during early post-stimulus intervals. This figure complements Figure 2, which presents results using windowed polynomial detrending (degree=3) for the same datasets (D5, D6, D7).

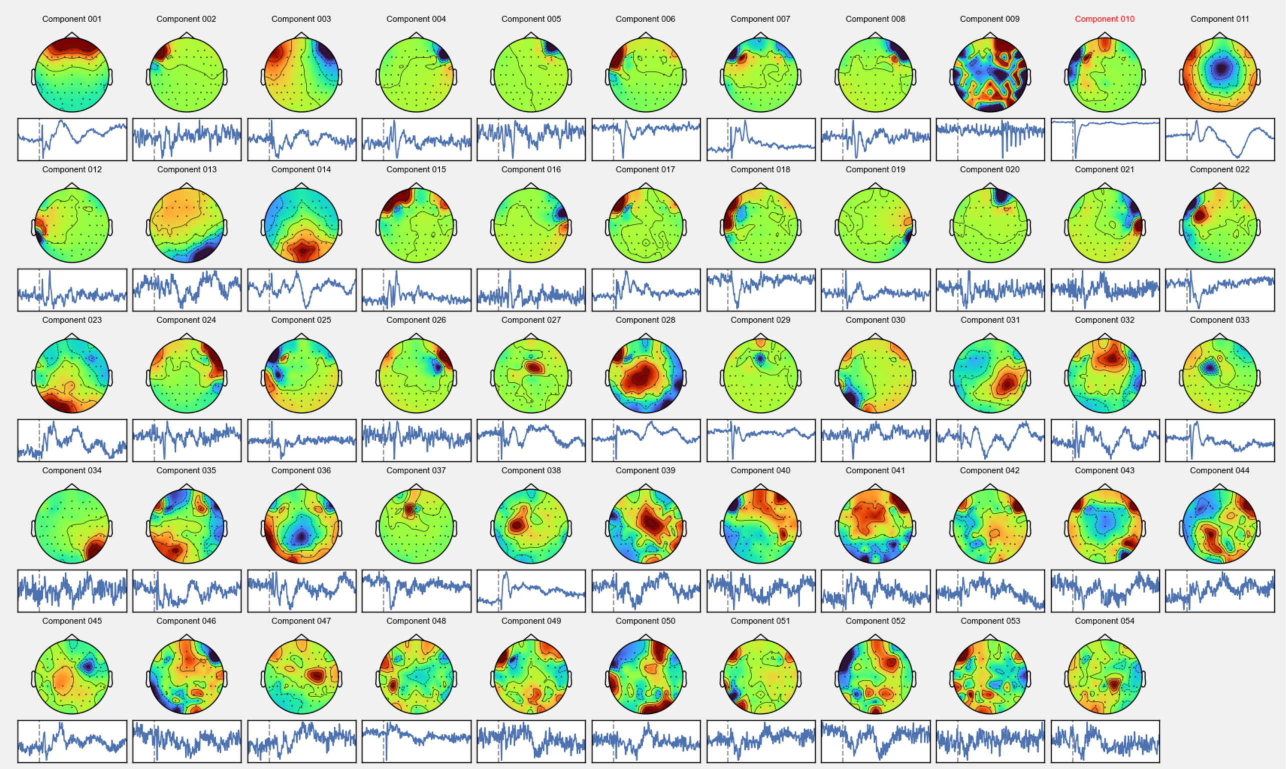


**Supporting Figure 4. ICA output components from dataset D5.** Components highlighted in red were excluded as they represent capacitive artifacts (rise + decay). The excluded component in red is number 010, that is a total of 1 out of 55 (1.8%), indicating a minimal presence of capacitive artifacts in this dataset. Dataset 5 is collected at the Chalfont Centre for Epilepsy (Buckinghamshire, UK).

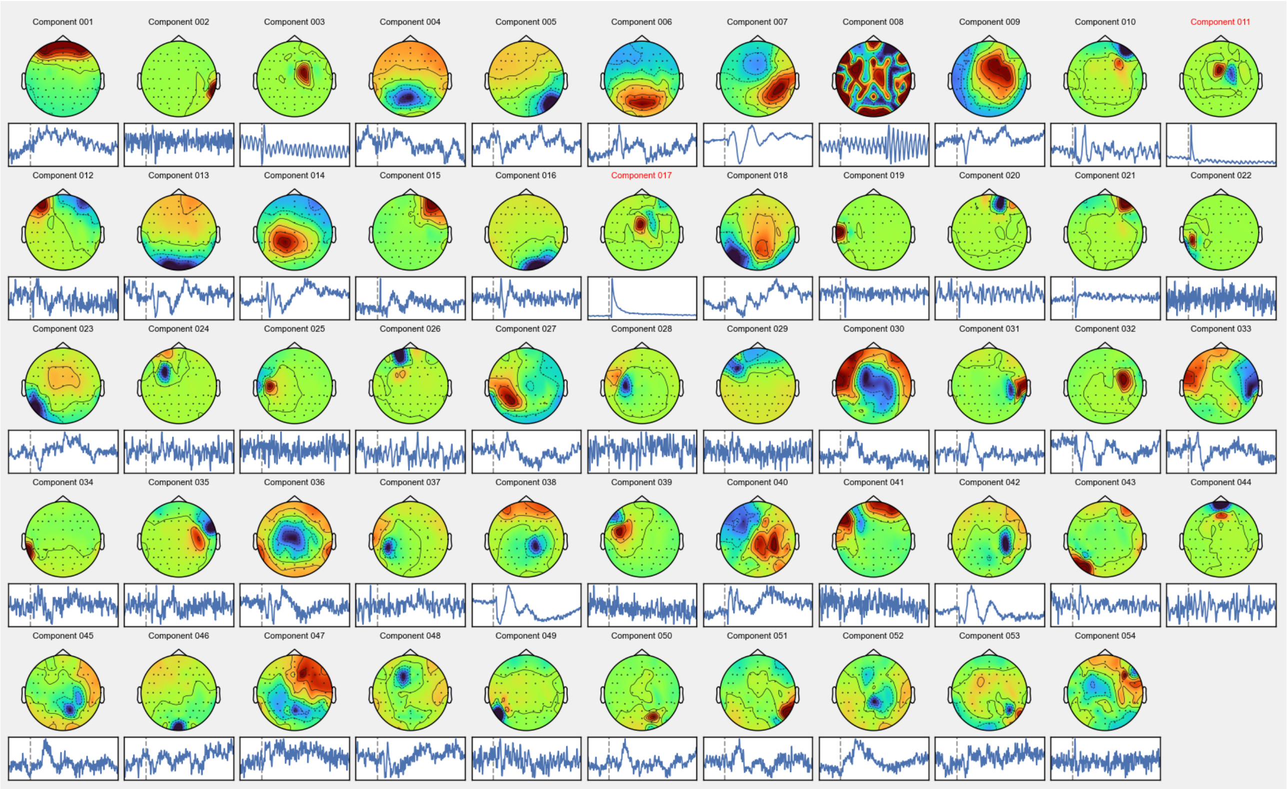


**Supporting Figure 5. ICA output components from dataset D6**. Components highlighted in red were excluded as they represent capacitive artifacts (rise + decay). The excluded components in red are numbers 011 and 017, that is a total of 2 out of 55 (3.6%), indicating a limited presence of capacitive artifacts in this dataset. Dataset 6 is collected at the Chalfont Centre for Epilepsy (Buckinghamshire, UK).

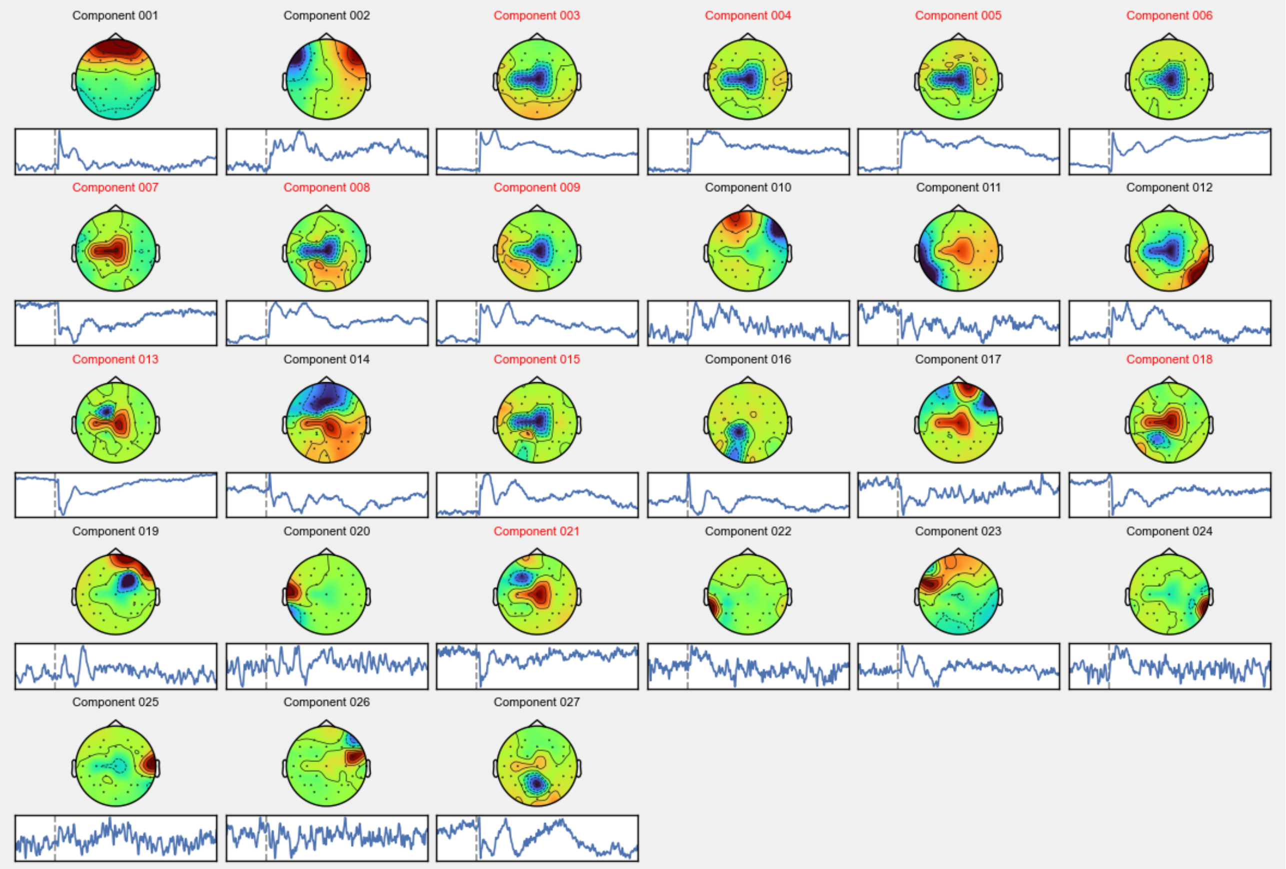


**Supporting Figure 6. ICA output components from dataset D7 (EBNeuroCap + BrainAmp + Nexstim).** Components highlighted in red were excluded as they represent capacitive artifacts (rise + decay). The excluded components in red are numbers 003, 004, 005, 006, 007, 008, 009, 013, 015, 018, 021, that is a total of 11 out of 27 (40.74%) components, demonstrating that extreme capacitive artifacts dominate the ICA analysis. In datasets with a limited number of channels, this dominance can compromise the extraction of other types of artifacts.

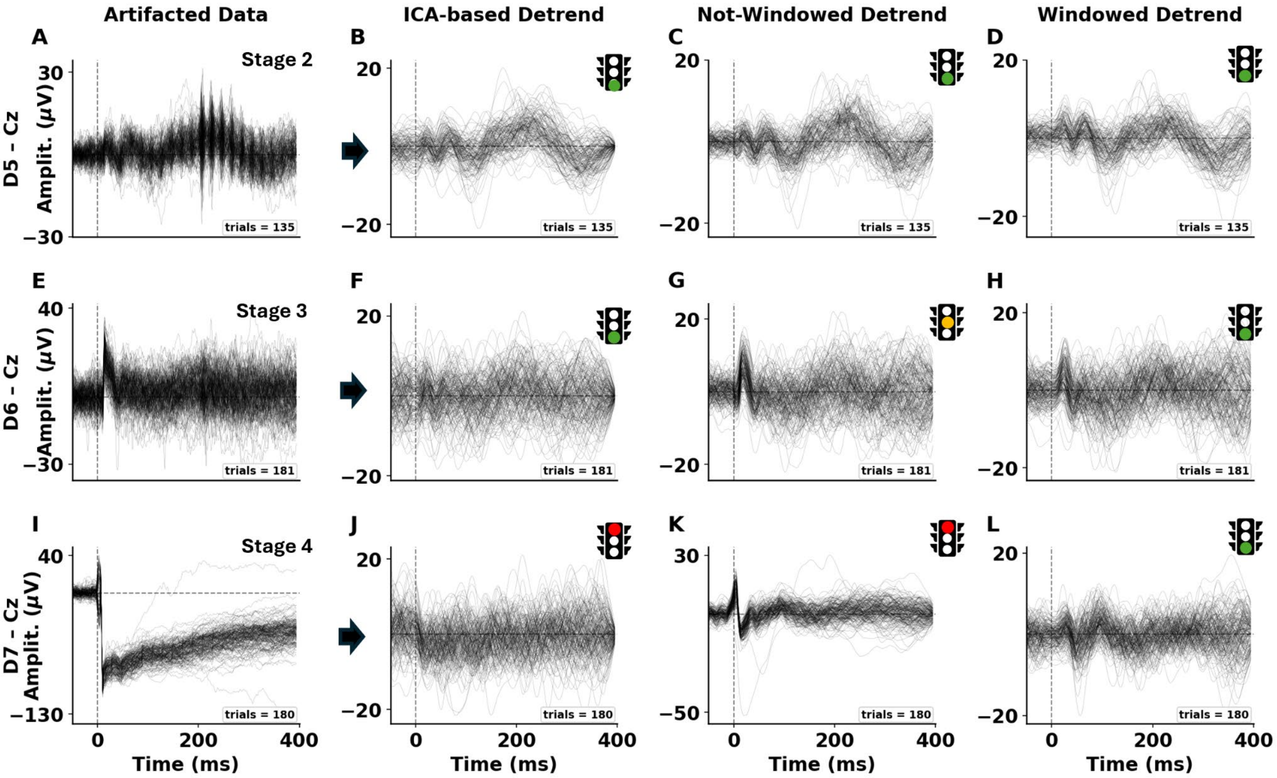


**Supporting Figure 7. Cz trial-wise waveforms across preprocessing methods.** Single-trial traces (gray) at channel Cz for each subject (rows: D5–D7) and preprocessing condition (columns: Artifacted, ICA-based, Non-Windowed, Windowed). The vertical dashed line marks *t* = 0 ms (TMS pulse); horizontal lines indicate 0 µV. Each panel shows all trials (n reported in the bottom-right inset). Datasets D5 and D6 were collected at the Chalfont Centre for Epilepsy (Buckinghamshire, UK) from already published work (D'Ambrosio et al., 2022), while D7 is the combination of EBNeuroCap + BrainAmp + Nexstim.

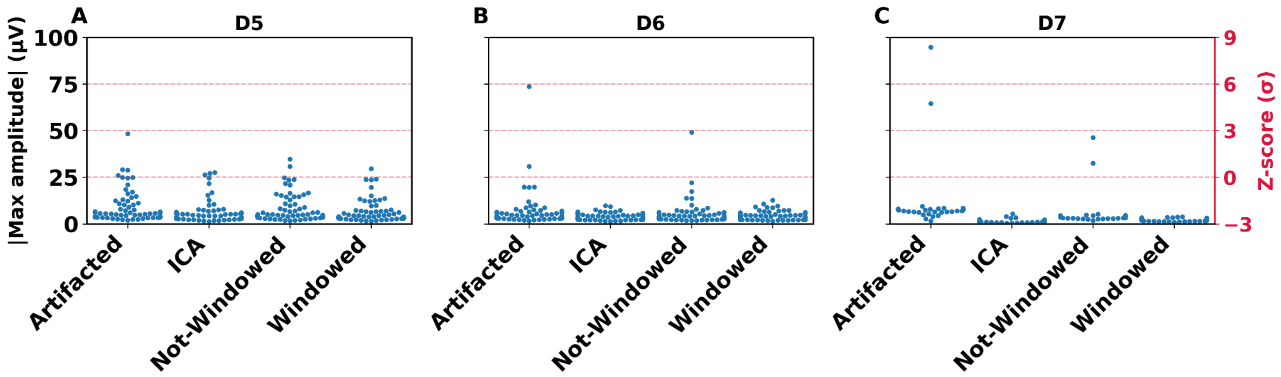


**Supporting figure 8. Peak amplitudes of averaged TEPs across preprocessing methods and datasets.** Each panel (A–C) shows the distribution of |max amplitude| (μV) across EEG channels for three subjects (D5–D7) and four conditions: Artifacted, and preprocessing methods: ICA, Non-Windowed, and Windowed. Blue dots represent channel-wise maximal TEP amplitudes; crimson dashed lines indicate corresponding z-score levels (–3σ to +9σ). The right y-axis shows standardized amplitude (Z-score), highlighting relative differences within each subject. The plots display the amplitude modulation by the preprocessing methods (refer also to Figure 2A/B). Datasets D5 and D6 are collected at the Chalfont Centre for Epilepsy (Buckinghamshire, UK) from already published work (D'Ambrosio et al., 2022), while D7 is the combination of EBNeuroCap + BrainAmp + Nexstim.

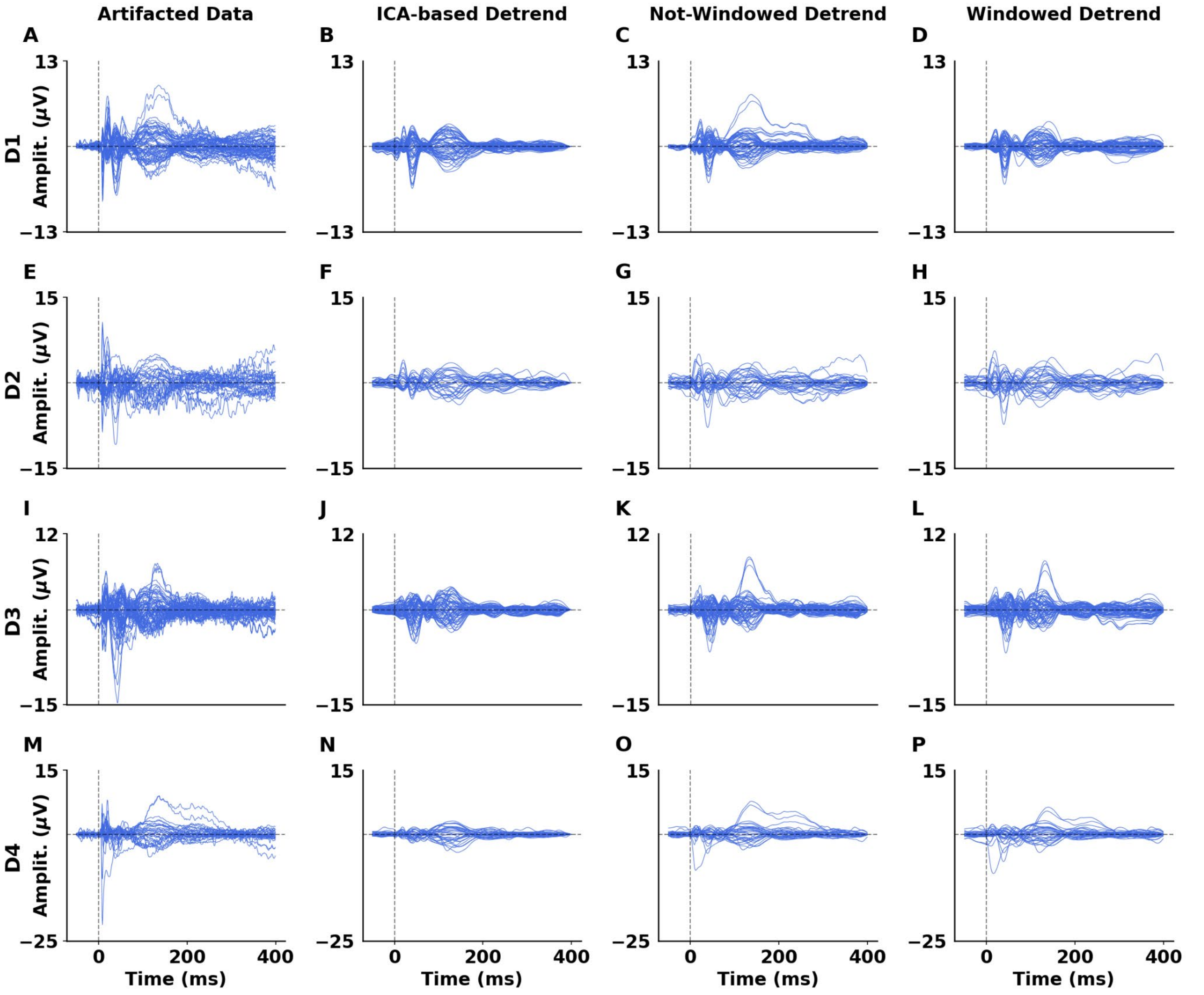


**Supporting Figure 9. Comparison of TEP waveforms after different artifact correction methods in D1/2/3/4 datasets.** Dataset 1, 2, 3, and 4 are without "over 3-sigma rule" amplitude deviation (see Figure 2A/B). Each panel shows single-trial signals (blue traces) from one subject and method. Columns correspond to preprocessing stages: **(A–D)** Subject D1, **(E–H)** D2, **(I–L)** D3, **(M–P)** D4. From left to right: *Artifacted Data*, *ICA-based Detrend*, *Non-Windowed Detrend*, and *Windowed Detrend*. Each trace represents the average across selected EEG channels (μV) time-locked to the TMS pulse (0 ms, dashed line). The combinations of hardwares of the datasets are D1 = EasyCap + BrainAmp + Nexstim; D2 = EasyCap + EBNeuro amplifier + Nexstim; D3 = EasyCap + BrainAmp + EBNeuroStim; D4 = EasyCap + EBNeuro amplifier + EBNeuroStim)

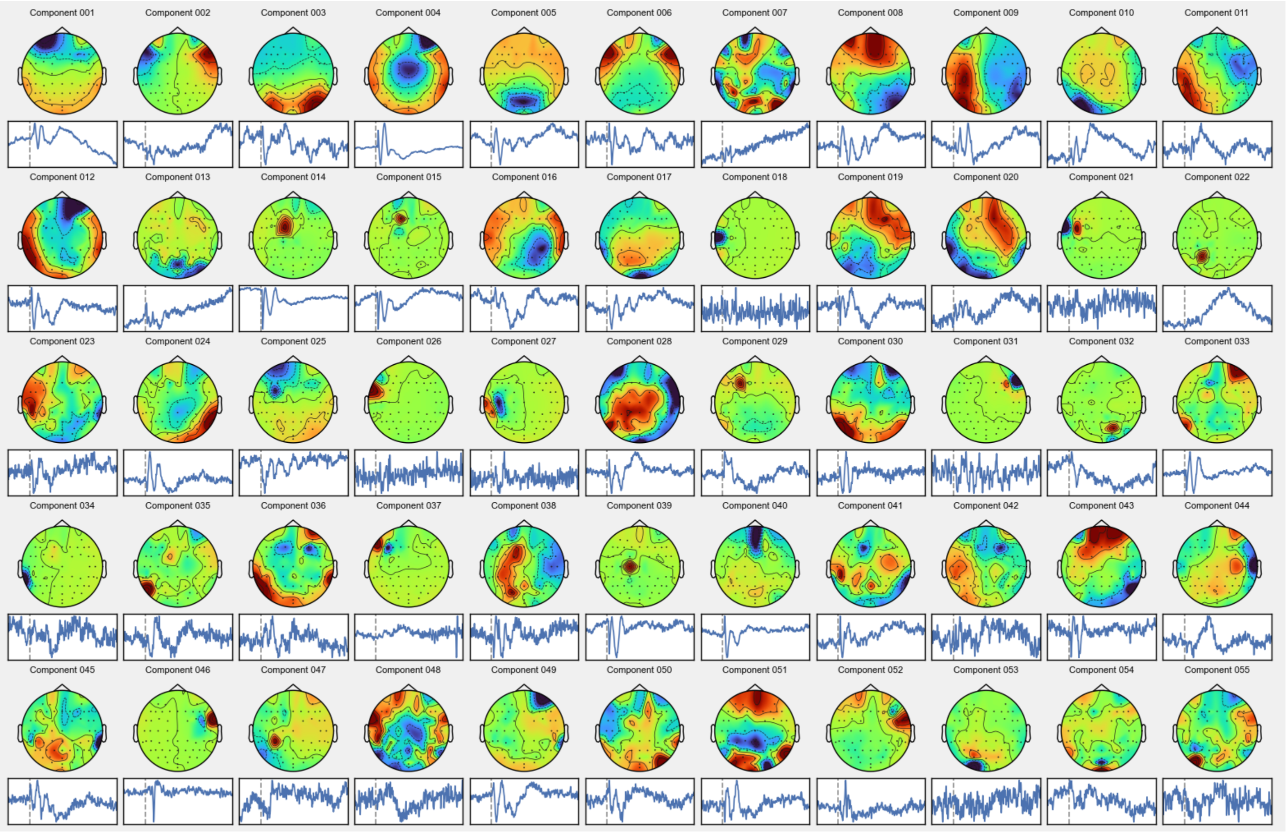


**Supporting Figure 10. ICA output components from dataset D1 (EasyCap + BrainAmp + Nexstim).** Components highlighted in red were excluded as they represent capacitive artifacts (rise + decay). In this dataset there are no capacitive artifact components.

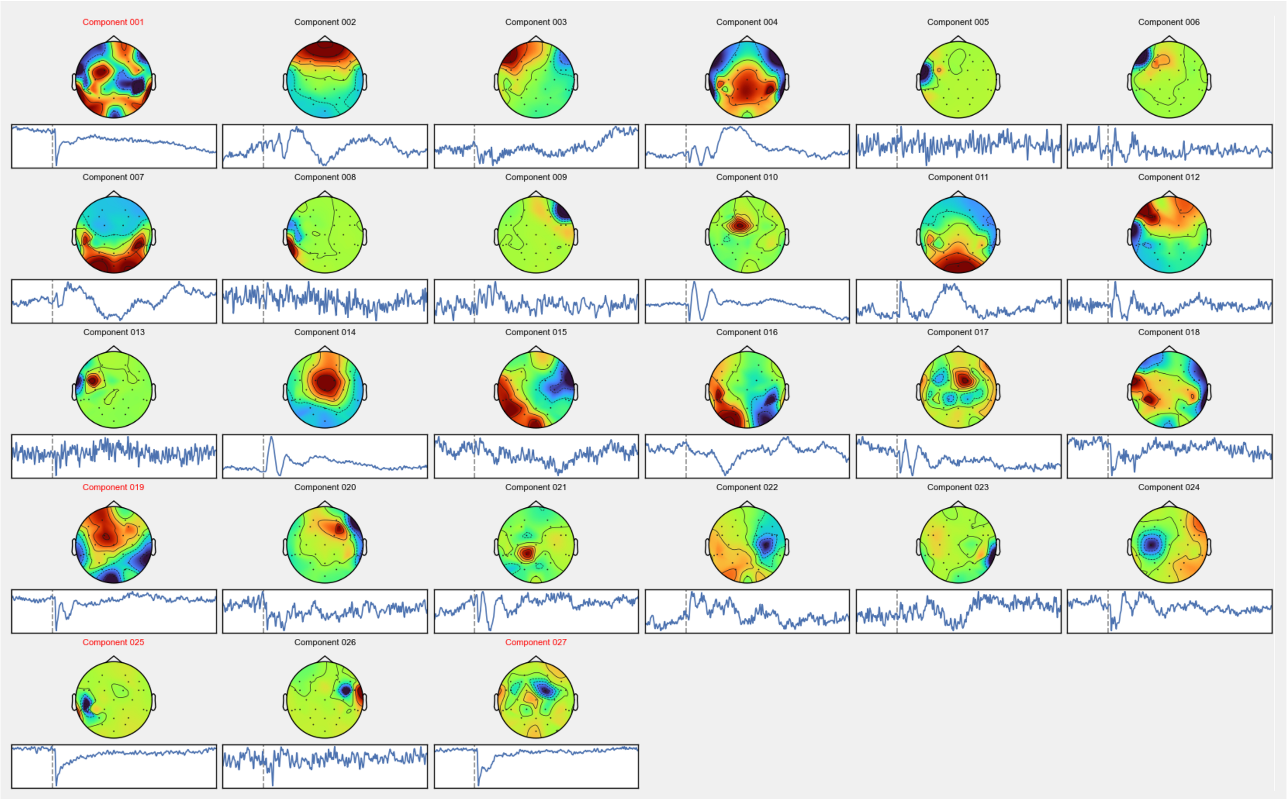


**Supporting Figure 11. ICA output components from dataset D2 (EasyCap + EBNeuro amplifier + Nexstim).** Components highlighted in red were excluded as they represent capacitive artifacts (rise + decay). The excluded components in red are numbers 001, 019, 025, 027, that is a total of 4 out of 27 (14.8%) components.

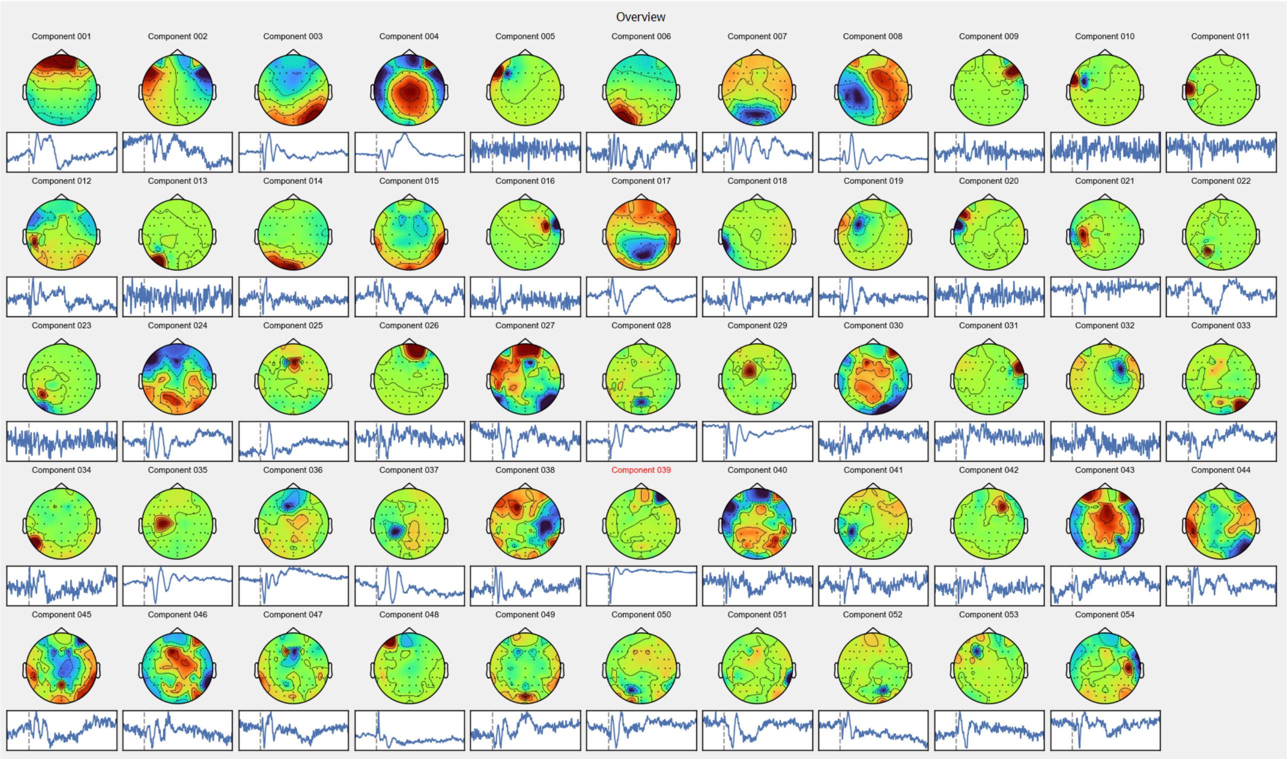


**Supporting Figure 12. ICA output components from dataset D3 (EasyCap + BrainAmp + EBNeuroStim).** Components highlighted in red were excluded as they represent capacitive artifacts (rise + decay). The excluded component in red is 039 that is a total of 1 out of 27 (3.7%) components.

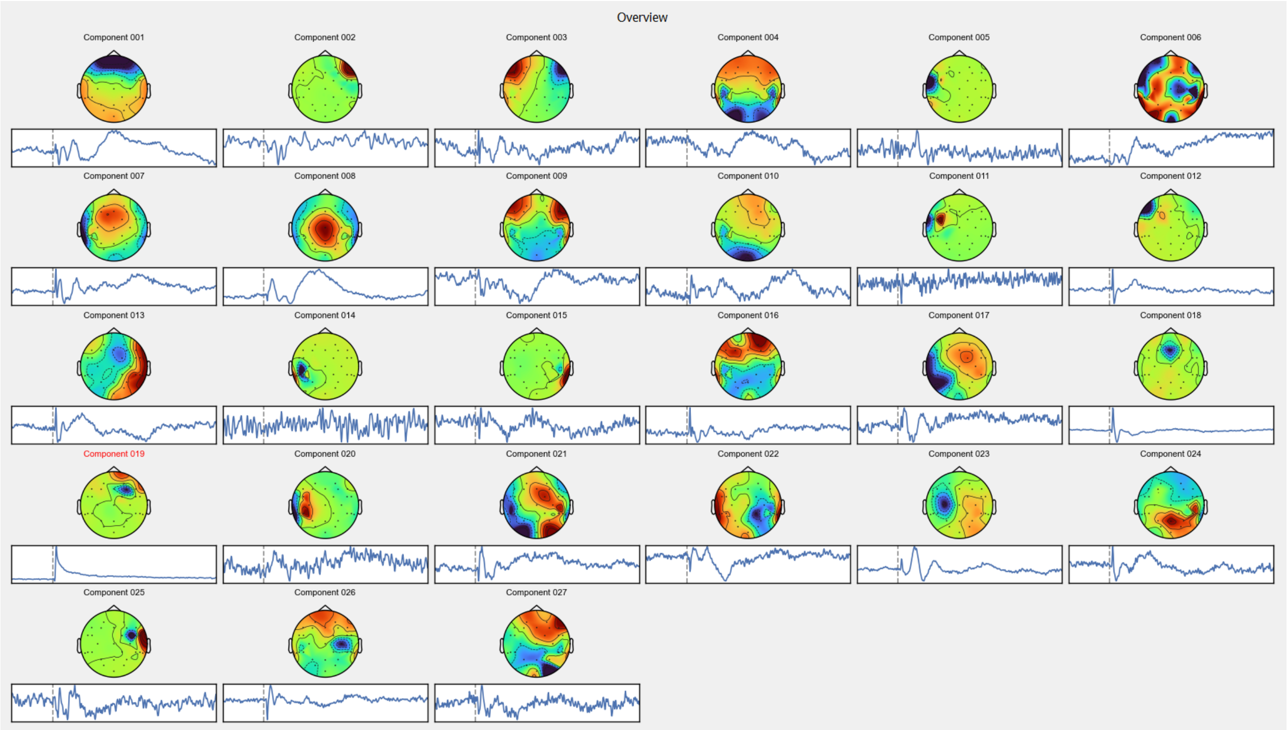


**Supporting Figure 13. ICA output components from dataset D4 (EasyCap + EBNeuro amplifier + EBNeuroStim).** Components highlighted in red were excluded as they represent capacitive artifacts (rise + decay). The excluded component in red is number 019, that is a total of 1 out of 27 (3.7%) components.
